# Medication Use in Severe Anorexia Nervosa: A Danish Register-based Study

**DOI:** 10.1101/2025.07.15.25330554

**Authors:** Zi-Ping Zhang, Hannah Chatwin, Janne Tidselbak Larsen, Loa Clausen, Esben Agerbo, Trine Munk-Olsen, Kathrine Bang Madsen, Bjarni Jóhann Vilhjálmsson, Liselotte Vogdrup Petersen, Zeynep Yilmaz

## Abstract

**Background:** Severe anorexia nervosa (AN) represents a subgroup of individuals with AN with prolonged illness duration and poor prognosis. Previous research has reported increased medication use in AN, but prescription patterns in severe AN remain unexplored.

**Objectives:** This study aimed to: (1) compare medication prescriptions between individuals with severe and less-severe AN; (2) explore the heterogeneity of prescription patterns among individuals with severe AN across different comorbidity profiles.

**Methods:** Utilizing Danish registers, this cohort study included 7,654 individuals diagnosed with AN. We assigned cases to severe or less-severe groups based on their AN Register-based Severity Index scores. First, we examined trajectories of medication prescriptions and compared patterns between groups using logistic regression models. Second, common comorbidity profiles among individuals with severe AN were identified by latent class analysis for further between-group comparisons. Sensitivity analyses were conducted to test alternative definitions of severe AN.

**Findings:** Compared to individuals with less-severe AN, those with severe AN were more likely to be prescribed various medications, including drugs targeting alimentary tract (OR = 1.4, 95% CI 1.3-1.6), cardiovascular drugs (OR = 1.1, 95% CI 1.0-1.3), analgesics (OR = 1.2, 95% CI 1.1-1.3), and psychotropic drugs (OR = 2.4, 95% CI 2.1-2.7). Notably, this pattern persisted even among individuals without diagnosed comorbidities. Within the severe group, five clusters with distinct comorbidity profiles emerged, and we consistently observed greater prescription rates than in the comorbidity-free cluster. Sensitivity analyses confirmed that our severity classification reliably distinguished severe from less-severe AN across multiple definitions.

**Conclusion:** These findings indicate that severe AN is associated with substantially higher prevalence of prescribed medications, while specific comorbidity patterns further influence prescribing patterns. This diverse and prolonged pharmacological treatment in severe AN reflects the complexity of clinical management in this population.

**Clinical implications:** Considering the widespread medication prescription in severe AN despite the lack of specific approved pharmacotherapy for the disorder, evidence-based treatment guidelines are urgently needed. Clinicians should recognize the substantial heterogeneity in severity and comorbidity burden within this population and develop comprehensive and specialized treatment strategies that evaluate risks and treatment needs across all individuals with AN, irrespective of comorbid conditions.

**Key messages:** *What is already known on this topic?:*

- The treatment of anorexia nervosa remains challenging due to the poorly understood etiology, compounded by the absence of approved pharmacotherapy and limited evidence for medication effectiveness.
- Individuals with anorexia nervosa and eating disorders in general receive multiple medications, but prescription patterns specifically in severe AN cases have not been systematically examined.

*What this study add?:*

- This study is the first to focus specifically on severe anorexia nervosa with long-term follow-up, with a precise definition of severity using the Anorexia Nervosa Register-based Severity Index in comprehensive Danish national registers.
- Individuals with severe AN have significantly higher prescription rates and more prolonged durations of pharmacotherapy compared to less-severe cases, with substantial prescribing observed even in the absence of diagnosed comorbidities.
- Within the severe anorexia nervosa population, individuals with distinct comorbidity profiles show associations with different medication patterns, providing a multidimensional understanding of pharmacological management.
- Timeframe selection critically influences the anorexia nervosa severity classification, as 5- or 7-year post-diagnosis periods provide more robust distinctions, while a shorter assessment period insufficiently captures the full picture of illness complexity.

*How this study might affect research, practice, or policy?:*

- This study provides essential evidence for the need to develop evidence-based pharmacotherapy guidelines for severe anorexia nervosa, addressing the current gap between extensive medication prescriptions and the lack of approved treatments.
- Clinicians should consider both illness severity and individual clinical presentations to optimize personalized treatment and address potential medication-related risks in this vulnerable population.

## INTRODUCTION

Anorexia nervosa (AN), a serious eating disorder (ED) affecting 4% of females and 0.3% of males worldwide ^[1]^, carries the risk of high chronicity. Follow-up studies have found that 20% of individuals still meet AN criteria five years post-treatment ^[2]^, and recovery rates remain below 80% even after 20 years ^[3]^, highlighting the critical need for effective treatments.

The treatment of AN is difficult, with limited evidence supporting pharmacological interventions.^[4]^ No medication has been approved by the U.S. Food and Drug Administration for AN or demonstrated consistent or extensive efficacy.^[5]^ Nevertheless, pharmacotherapy is commonly utilized for individuals with AN ^[5]^, often targeting specific symptoms rather than the core pathology. More recently, a Danish population-based study reported higher prescription rates of any type of medication in AN patients compared to population-matched controls.^[6]^ However, existing research rarely distinguishes severe and less-severe AN, leaving pharmacotherapy patterns specific to severe AN poorly understood.

The challenge in treating AN is complicated by the high prevalence of comorbidities. Compared to other mental disorders, AN demonstrates notably higher psychiatric comorbidity rates, ranging from 58% to 87% of individuals with AN having a comorbid psychiatric disorder during their lifetime.^[7]^ Common psychiatric comorbidities include mood disorders, anxiety disorders, obsessive-compulsive disorder, substance use disorders, and personality disorders. Despite their typical association with advanced age, elevated risks of somatic illness ^[8]^, including but not limited to cardiovascular complications, gastrointestinal diseases, osteopenia, and osteoporosis, have been reported in relatively young individuals with AN. Consequently, treatment objectives typically address both psychopathology and associated medical conditions, leading to considerable variations in medication prescription patterns, particularly across comorbidity profiles and illness severity.

The exploration of pharmacotherapy patterns in severe AN is impeded by the lack of clinical criteria or definition of severity. The recently developed AN Register-based Severity Index (AN-RSI) provides a unique opportunity to distinguish severe cases from the broader AN population in the absence of detailed clinical notes.^[9]^ Leveraging data from population registers, AN-RSI is designed to measure AN severity as continuous scores, incorporating multiple register-based indicators that align with key elements of the severity guidelines ^[10, 11]^, ensuring more accurate classification of individuals.

Considering the complexities associated with AN treatment, this study aimed to examine prescription patterns in severe AN with two distinct objectives. We first compared medication prescriptions between severe and less-severe AN groups, hypothesizing that severe AN cases would demonstrate significantly higher use of both psychotropic and somatic medications. Additionally, we investigated the association between comorbidity profiles and prescription patterns in severe AN. Specifically, we hypothesized that distinct comorbidity profiles would correlate with varying medication regimens, with certain comorbidity patterns associated with more frequent and diverse medication prescriptions.

## METHODS

### Study design

This register-based cohort study comprised all individuals diagnosed with AN (ICD-8: 306.50; ICD-10: F50.0-50.1) in Denmark between 1 January 1991 and 31 December 2011. Cases were identified through the Danish National Patient Register (DNPR) and the Danish Psychiatric Central Research Register (DPCRR).^[12, 13]^ Exclusion criteria included age younger than 6 years at first AN diagnosis, death, or emigration within 5 years after first AN diagnosis.

We utilized the existing severity scores calculated using AN-RSI at five years after initial diagnosis, incorporating the weight specified in the Supplementary Table S1 (for details, see Larsen *et al*.^[9]^). In the primary analysis, severity assessment was made at 5 years after initial diagnosis. Patients who died or emigrated within 5-10 years after the first AN diagnosis were censored from the analysis at the time of death or emigration. Due to the differences of opinion in the field of EDs regarding the illness duration criterion of severe AN, there is no agreed-upon minimum threshold in the literature. Many prior studies set the duration threshold to either 3 or 7 years ^[11, 14]^, and in order to balance adequate capture of illness progress, we selected an intermediate 5-year duration as our threshold. Individuals scoring in the top 20% on AN-RSI were categorized as severe cases, with the remaining individuals classified as less-severe. This threshold aligns with previous studies demonstrating that approximately 20% of individuals with AN have poor treatment outcomes ^[15]^, supporting the clinical and evidence-based relevance of this cutoff for identifying severe cases.

Prescription records between 1996 and 2021 were obtained from the Danish National Prescription Registry ^[16]^, which includes prescriptions redeemed from general practitioners and community pharmacies in Denmark. This period was selected to ensure a full five-year follow-up period after establishing AN severity. Unique personal identification numbers from the Danish Civil Registration System facilitated data linkage across nationwide health registers.^[17]^ To maintain consistency with prescription registries, medications were grouped according to the Anatomical Therapeutic Chemical classification codes.^[18]^ Primary classes of medication included drugs for the alimentary tract (A), cardiovascular drugs (B, C), anti-infective drugs (D, J, P), genitourinary system and sex hormones (G), hormonal preparations (H), immunomodulating drugs (I), musculoskeletal drugs (M), analgesics (N02), psychotropic drugs (N, excluding N02), respiratory and sensory drugs (R, S), and others (V).

Subclasses of medication categories and specific medications of interest were examined separately, including those detailed in the Supplementary Table S2. Medication prescription was treated as a binary variable and defined by the redemption of at least two prescriptions within a given medication group during the observation period.

A set of diagnoses from the ICD, including twelve somatic chapters and twelve psychiatric chapters, was extracted from Danish health registers and coded as either present or absent. The rationale for the selection of chapters and specific definitions used was detailed in Supplementary Table S3.

### Statistical analyses

At the 5-year post-diagnosis point, we classified patients as having severe or less-severe AN based on AN-RSI scores and reported the onset age and sex distributions of both groups. We explored prescription trajectories for these severity groups both retrospectively (from the initial diagnosis to the 5-year classification point) and prospectively (from classification to 10 years post-diagnosis). To prospectively examine whether severity classification at the 5-year time point is associated with subsequent prescription patterns, we quantified differences between the two groups and calculated odds ratios (ORs) with 95% confidence intervals (CIs) for prescriptions during the period spanning from five to ten years post-diagnosis, thereby avoiding comparing groups before the severity classification. To delineate the effects of AN severity, we examined prescription patterns over 5–10 years post-diagnosis in patients without psychiatric and somatic comorbidities. All models for these analyses were adjusted for calendar year to reduce temporal bias.

Statistical clustering techniques such as latent class analysis (LCA) were applied to explore the heterogeneity of diseases and comorbidities. With the concurrent psychiatric and somatic diagnoses as input features, we performed LCA to identify subgroups with different levels of comorbidity burden in severe AN. Briefly, severe AN cases without any comorbidities were assigned to a separate cluster and excluded from LCA. Models with two to six clusters were assessed, with the optimal model selected based on statistical measures (Supplementary Table S4). Individuals were assigned to clusters according to their highest probability of membership. K-means was employed to validate the robustness of LCA results. ORs for differences in prescription rates across medication classes were compared between groups with and without comorbidities, with p-values adjusted via Benjamini-Hochberg correction. Further comparisons were stratified by comorbidity profile clusters.

### Rationale for sensitivity analyses

In the primary analysis, severity assessment was made at 5 years after initial diagnosis. Sensitivity analyses employed the alternative timeframes – 3-and 7-year post-diagnosis - to establish the AN severity and further examine the association with medication prescription. These timeframes were selected to address ongoing discourse in the field regarding optimal illness duration criteria for defining severe AN, ranging from 3 to 7 years or longer ^[11, 14]^, and to validate our findings across different definitions. Prescription patterns from 5 to 10 years or 7 to 10 years post-diagnosis were compared between severe and less-severe groups under each severity classification (Supplementary Figure S1). LCA identified comorbidity profiles among severe AN patients defined by AN-RSI at these alternative timepoints (versus the original 5-year AN-RSI).

## RESULTS

### Population characteristics

A total of 7,654 individuals were included in the main analyses, with 1,652 (21.6%) classified as severe AN cases based on AN-RSI (Supplementary Table S5). The severe AN group had a slightly higher proportion of females (95.2% vs. 93.2%, p=0.004) and similar age at AN diagnosis (19.6 vs. 19.7 years, p=0.481). Individuals with severe AN presented significantly more frequently with any comorbidity (89.8% vs. 81.3%, p<0.001), including both psychiatric (71.1% vs. 53.6%, p<0.001) and somatic conditions (74.1% vs. 66.5%, p<0.001).

### Severe AN versus less-severe AN

The trajectories of medication prescription among individuals with severe AN and less-severe AN are shown in Figure 1, capturing prescriptions over 10 years after first AN diagnosis (5 years before and 5 years after the severity assessment). Medication for somatic illnesses stabilized during follow-up for both groups. However, medications for the genitourinary system and sex hormones peaked later in the severe AN group, at 8 years after the first AN diagnosis, compared to 5 years in the less-severe AN group. Psychotropic drug prescriptions were consistently higher in severe AN, primarily driven by the difference in antidepressants, antipsychotics, antiepileptics, and anxiolytics/hypnotics (Figure 1c).

**Figure 1.**
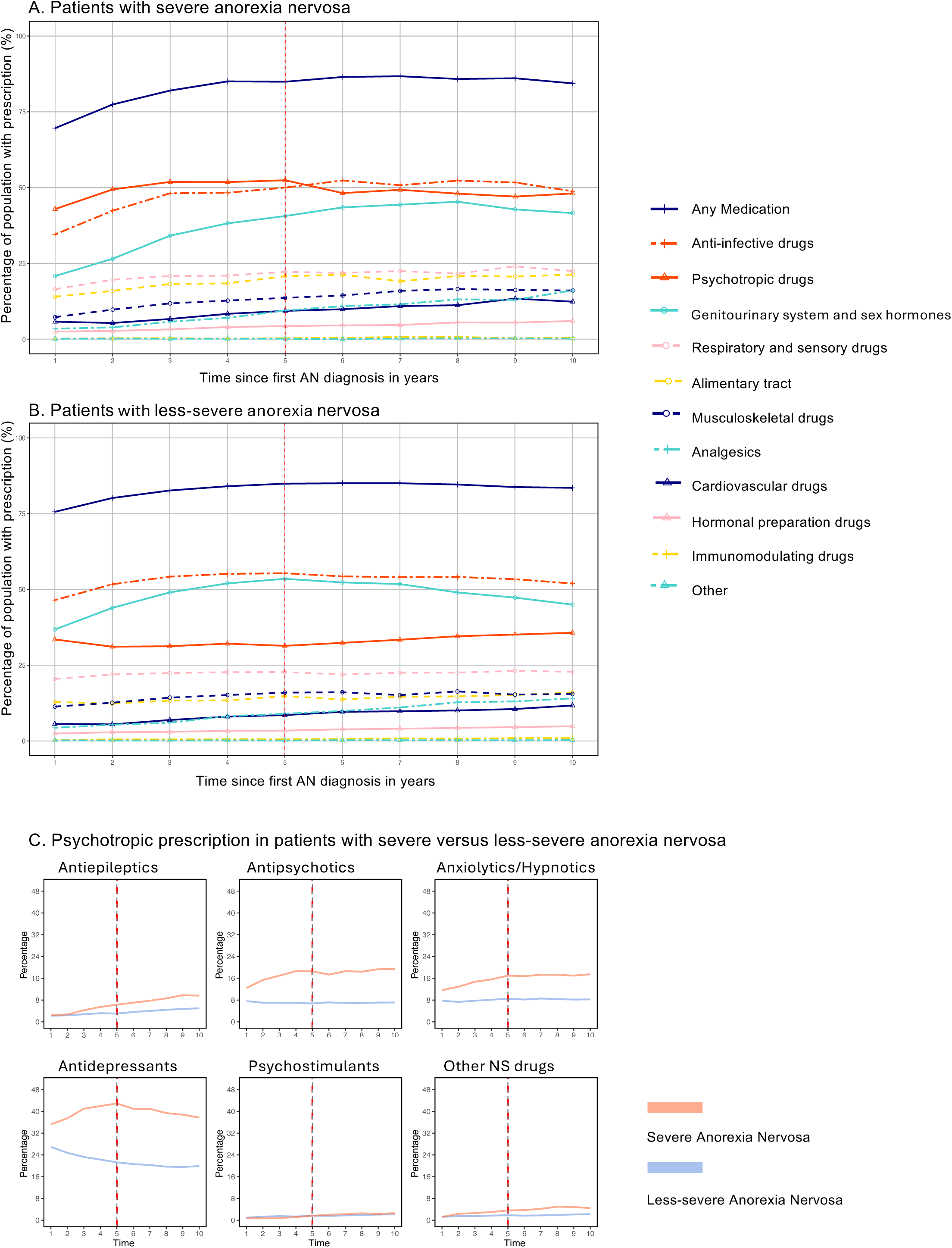
Trajectory of medication prescriptions within 10 years post-diagnosis. Medication prescription trajectories were restricted to individuals diagnosed with AN between 1996 and 2011 to ensure complete 10-year prescription coverage and to avoid left-censoring. The corresponding sample included 5,437 individuals (severe AN: n = 1,227; less-severe AN: n = 4,210). The y-axis shows the percentage of patients with prescriptions, calculated as the proportion who redeemed at least two prescriptions within discrete one-year intervals following their first AN diagnosis. The x-axis represents years since the first AN diagnosis. The red line represents the year when the Anorexia Nervosa Register-based Severity Index was used to assess illness severity. A. Medication prescription patterns over time in patients with severe anorexia nervosa (n = 1,652). B. Medication prescription patterns in patients with less-severe anorexia nervosa (n = 6,002). Overall prescription rates were lower compared to severe cases, particularly for psychotropic medications. C. Comparison of specific psychotropic medication classes between severity groups. Higher prescription rates were observed in severe anorexia nervosa across most categories. Other NS drugs: other nervous system drugs.

Figure 2 presents the ORs of prescriptions in severe AN compared to less-severe cases 5 years after establishing severity. Individuals with severe AN had higher likelihood of receiving medications for the alimentary tract (OR = 1.44, 95% CI 1.29-1.60), cardiovascular drugs (OR = 1.12, 95% CI 0.98-1.26), analgesics (OR = 1.18, 95% CI 1.05-1.32), and psychotropic drugs (OR = 2.39, 95% CI 2.11-2.71). Conversely, drugs for the genitourinary system and sex hormones, including sex hormones, were prescribed less frequently to individuals with severe AN, with ORs of 0.69 (95% CI 0.61-0.78) and 0.79 (95% CI 0.70-0.88), respectively. Within the psychotropic category, distinct prescription patterns for specific drugs were observed (Supplementary Figure S2). Atypical agents showed higher prescription rates compared to typical antipsychotics in both severity groups, as did selective serotonin reuptake inhibitors (SSRIs) compared to non-selective serotonin reuptake inhibitors (non-SSRIs). Quetiapine was the most frequently prescribed antipsychotic in both groups (13.1% and 6.1%), followed by chlorprothixene (10.7% and 4.1%). However, the highest OR was observed for olanzapine use (3.74; 95% CI 2.92-4.79). Sertraline was the most commonly prescribed SSRI (20.7% and 10.6%) and also exhibited the highest OR among SSRIs (OR = 2.21, 95% CI 1.91–2.55), followed by fluoxetine, which had the second-highest prescription rate and a similarly elevated OR (12.3% and 16.1%; OR = 2.17, 95% CI 1.81-2.60). Two non-SSRI antidepressants, mirtazapine and venlafaxine, were also frequently prescribed more in severe AN, with ORs of 1.63 (95% CI 1.33-1.99) and 1.84 (95% CI 1.51-2.25), respectively.

**Figure 2.**
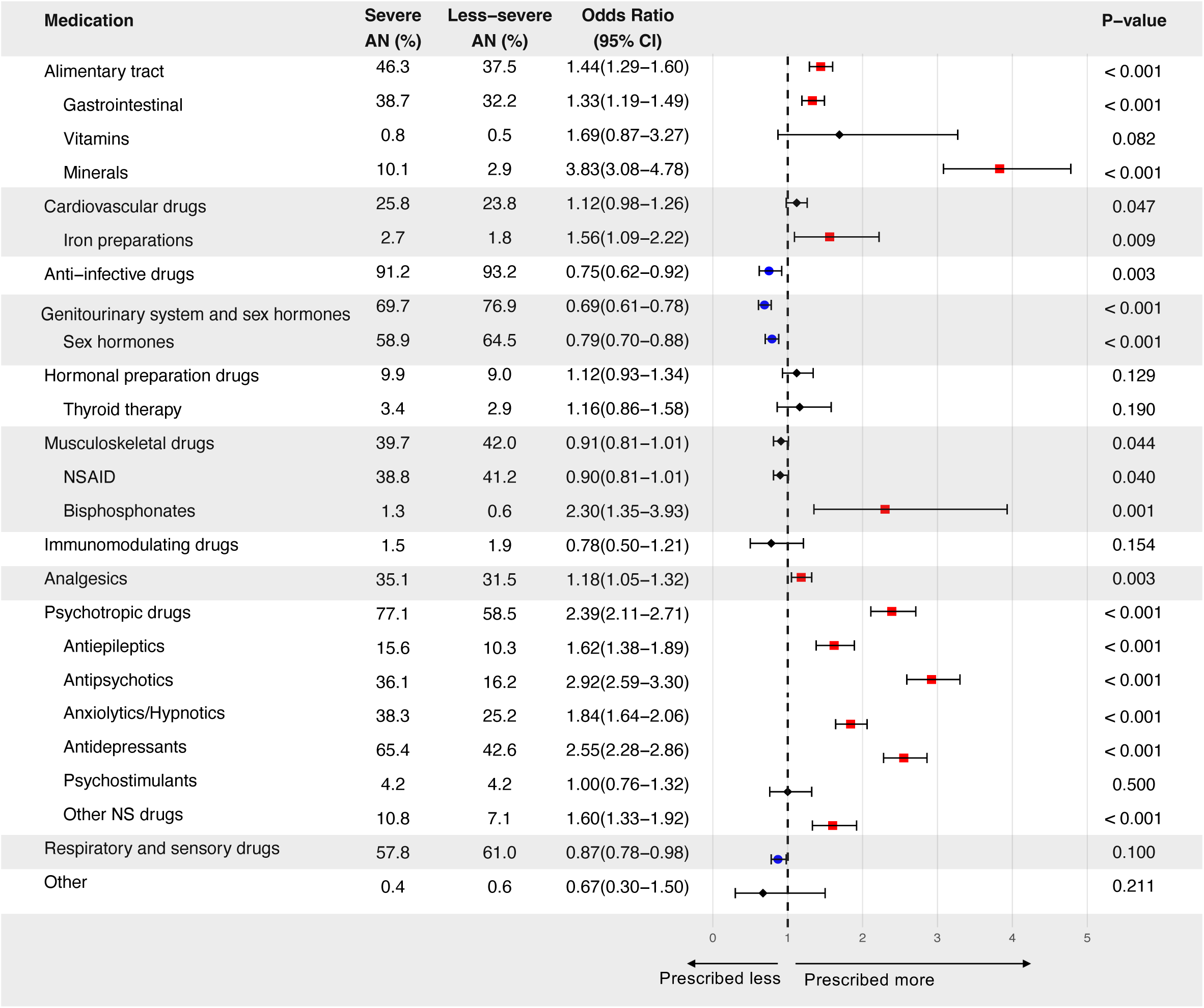
Medication prescriptions in patients with severe vs less-severe anorexia nervosa between 5 and 10 years post-diagnosis. This forest plot presents odds ratios (ORs) comparing medication prescription rates between severity groups during the 5-to 10-year follow-up period. ORs greater than 1.0 indicate higher prescription rates in severe AN. Horizontal lines represent 95% confidence intervals (CIs). AN, anorexia nervosa; NSAID, nonsteroidal anti-inflammatory drugs; Other NS drugs, other nervous system drugs.

When we examined AN cases without comorbidities (n = 169, representing 10.2% of severe AN cases; n = 1122, representing 18.7% of less-severe AN cases), both groups were prescribed a range of medications (Supplementary Figure S3). The highest prevalence was observed for anti-infective drugs in both groups, with 85.2% in the severe AN group and 87.6% in the less-severe AN group, although the difference was not statistically significant. Compared to less-severe AN cases, severe AN cases received more prescriptions for alimentary tract medications (OR = 1.41, 95% CI 0.97-2.05) and psychotropic drugs (OR = 2.40, 95% CI 1.72-3.33), particularly antidepressants (OR = 3.36, 95% CI 2.39-4.74) and antipsychotics (OR = 2.81, 95% CI 1.27-6.22). Medications for somatic conditions–except for vitamins/minerals–showed no significant variation between groups.

### Comorbidity profiles and medications in severe AN

Among the 1652 individuals with severe AN, 1483 (89.8%) had at least one additional diagnosis within the specified study timeframe, while the remaining 169 patients (10.2%) had no psychiatric or somatic diagnoses. Overall, 71.1% of individuals with severe AN had psychiatric comorbidities, and 74.1% had somatic comorbidities (Supplementary Table S5).

LCA identified clusters with differing clinical presentation in severe AN (Supplementary Figure S4a). Individuals without comorbidities were assigned to Cluster 1. Cluster 2 (27.8%) comprised AN cases with a low prevalence (< 30%) of both somatic and psychiatric comorbidities. Cluster 3 (42.2%) represented individuals with a medium burden (30–60%) of personality disorders, mood disorders, and anxiety disorders. Cluster 4 (10.2%) was characterized by high proportions (> 60%) of individuals with comorbid personality disorders, anxiety disorders, and self-harm, with moderate prevalence of mood disorders, schizophrenia, and substance use disorders. Individuals assigned to Cluster 5 (9.6%) exhibited high burdens (> 60%) of both psychiatric disorders and somatic illnesses. K-means clustering with 4-item–matching the optimal number of clusters from LCA–generated similar patterns (Supplementary Figure S4b).

Individuals with severe AN were prescribed various medications regardless of comorbidity burden (Table 1). Anti-infective drugs were most commonly prescribed (85.2-96.9% across all clusters). Prescription rates of medications for somatic conditions significantly differed among clusters based on somatic comorbidity burden. Psychotropic drugs were also prescribed frequently, ranging from 47.4% in individuals without comorbidity to over 90% in individuals with a high prevalence of psychiatric disorders (Clusters 4 and 5). Antidepressants were the most prevalent psychotropics, ranging from 41.4 % to 88.1%, followed by anxiolytics/hypnotics (10.1-77.4%) and antipsychotics (5.3-80.4%).

**Table 1.** Medication prescriptions by comorbidity profiles in patients with severe anorexia nervosa.

| Characteristics | Without comorbidity<br>N = 169 | Low burden of comorbidity<br>N = 459 | Medium burden of personality disorders, mood and anxiety disorders<br>N = 697 | High burden of psychiatric disorders and self-harm condition<br>N = 168 | High burden of comorbidity<br>N = 159 |
| --- | --- | --- | --- | --- | --- |
| Age at AN diagnosis (years) | 19.6 (6.2) | 19.6 (6.7) | 20.1 (6.7) | 19.9 (6.4) | 19.9 (6.9) |
| Alimentary tract | 25.4 | 41.4 | 40.8 | 65.5 | 86.8 |
| Gastrointestinal | 20.7 | 33.1 | 32.6 | 58.9 | 79.9 |
| Vitamins/Minerals | 5.9 | 11.1 | 12.9 | 20.2 | 29.6 |
| Cardiovascular drugs | 11.8 | 24.8 | 23.3 | 29.8 | 50.3 |
| Anti-infective drugs | 85.2 | 92.8 | 90.0 | 92.9 | 96.9 |
| Genitourinary system and sex hormones | 65.1 | 71.9 | 68.9 | 67.3 | 74.2 |
| Hormonal preparation drugs | 3.0 | 11.6 | 8.0 | 6.0 | 25.2 |
| Musculoskeletal drugs | 19.5 | 38.3 | 36.0 | 51.2 | 68.6 |
| Analgesics | 13.0 | 35.1 | 29.0 | 45.2 | 74.8 |
| Psychotropic drugs | 47.3 | 65.8 | 83.2 | 93.5 | 97.5 |
| Antidepressants | 41.4 | 47.1 | 72.9 | 88.1 | 87.4 |
| Anxiolytics/Hypnotics | 10.1 | 22.7 | 38.5 | 71.4 | 77.4 |
| Antipsychotics | 5.3 | 12.8 | 38.5 | 80.4 | 78.6 |
| Respiratory and sensory drugs | 40.8 | 60.4 | 53.2 | 62.5 | 83.7 |

Compared to those without comorbidities (Cluster 1), AN cases with varying degrees of comorbidity burden (Cluster 2-5) revealed significantly higher odds of receiving all medication classes except drugs for the genitourinary system and sex hormones (Figure 3). Notably, the prescription of psychotropic drugs was 43 times higher in individuals with a high burden of comorbidity (Cluster 5) compared to those without comorbidities. Medication prescriptions for the alimentary tract had approximately two-fold higher odds in low-burden clusters and nearly twenty-fold higher odds in patients with severe somatic disease burden. All pairwise comparisons between Cluster 1, 2, 3, and 4 also demonstrated similar trends: clusters with higher comorbidity burden had significantly elevated odds for most primary medication classes (Supplementary Figure S5)

**Figure 3.**
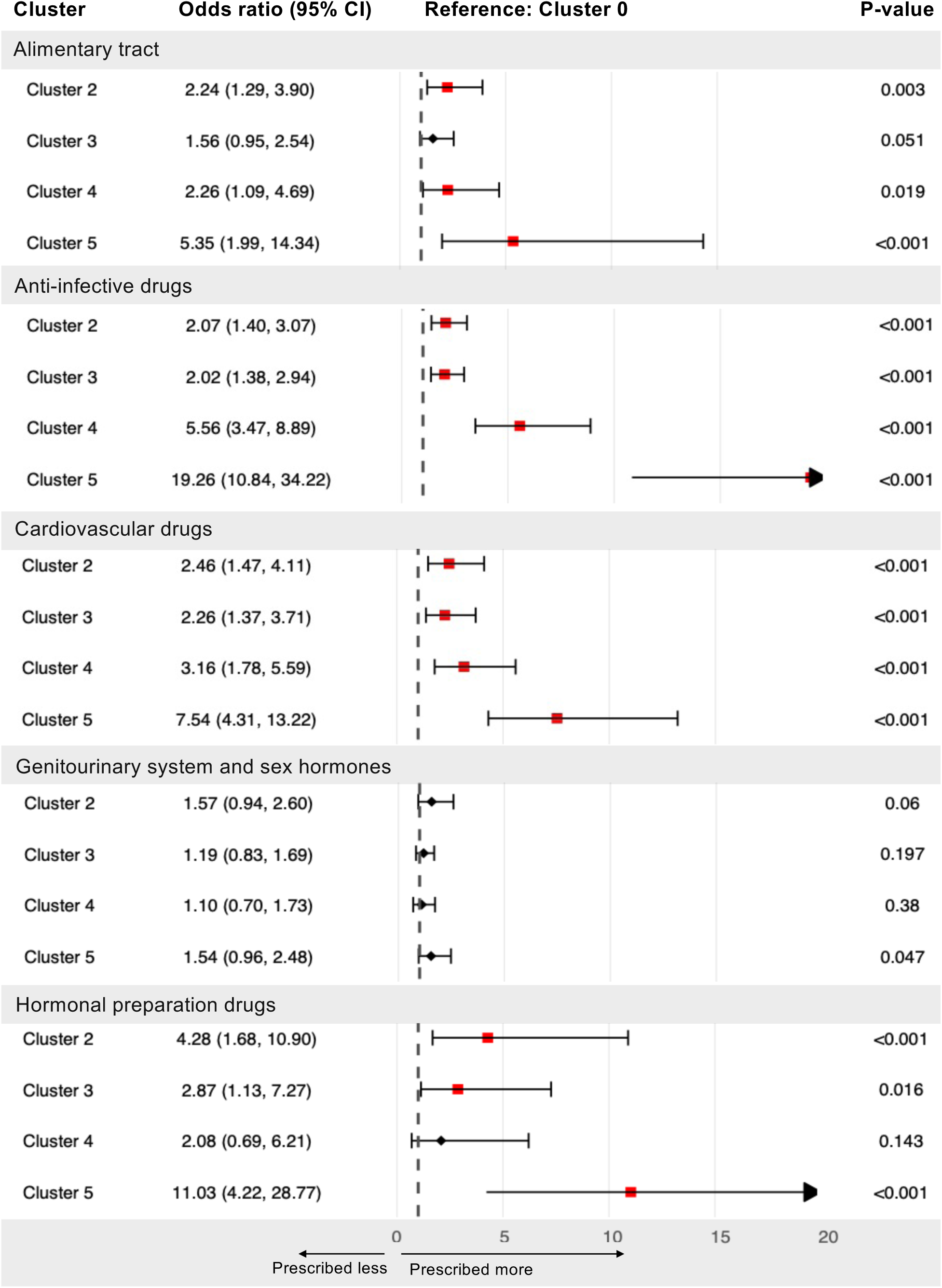
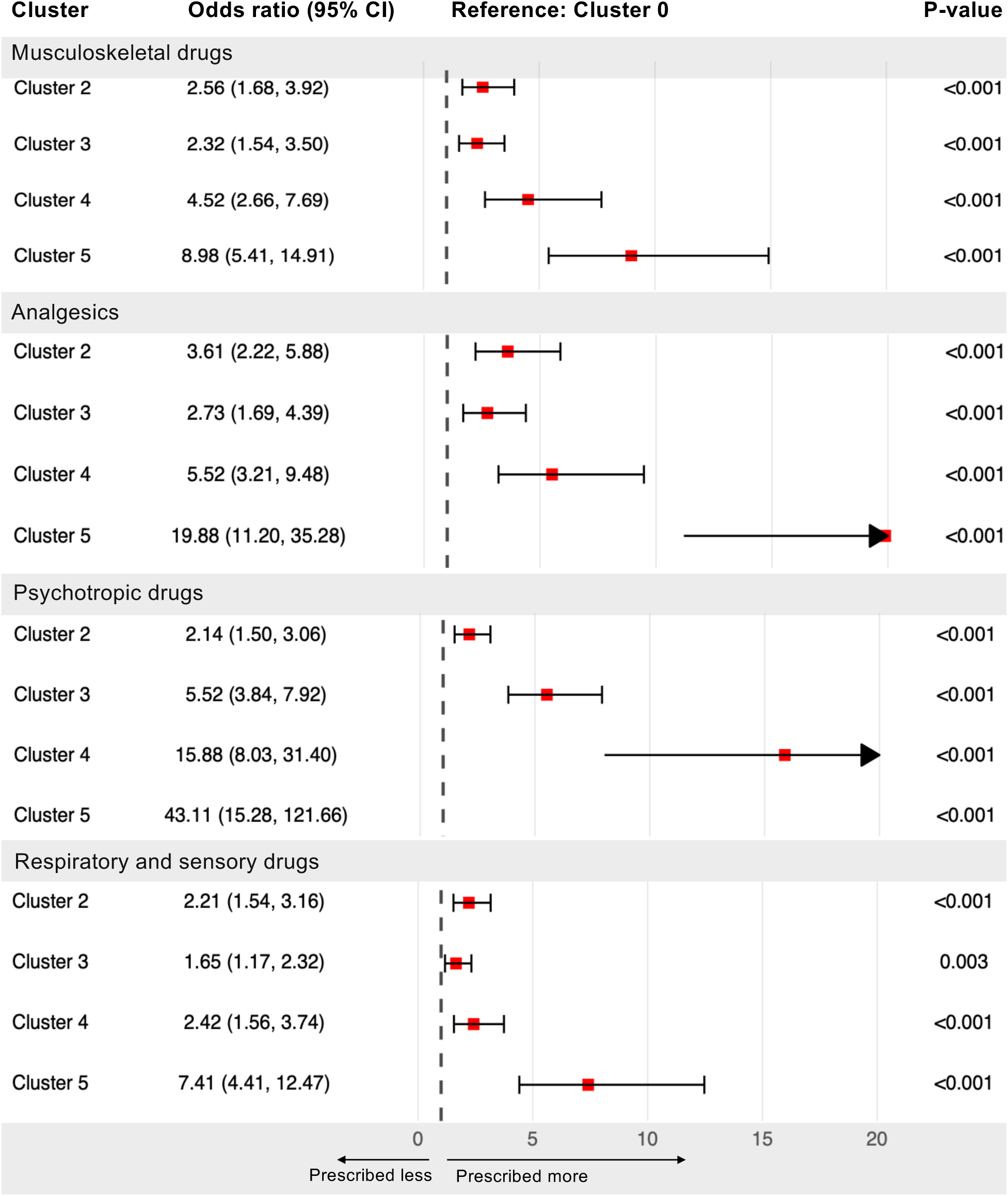
Medication prescriptions in severe anorexia nervosa patients with distinct comorbidity profiles. This forest plot presents odds ratios (ORs) comparing medication prescription rates between severe anorexia nervosa patients without comorbidity and the other four severe groups with distinct comorbidity profiles during the 5-to 10-year follow-up period. ORs greater than 1.0 indicate higher prescription rates in severe AN. Horizontal lines represent 95% confidence intervals (CIs). Cluster 1, without any comorbidities; Cluster 2: with low burden of comorbidities; Cluster 3: with a medium burden of personality, mood, and self-harm conditions; Cluster 4: with a high burden of personality disorders and self-harm conditions; Cluster 5: with a high burden of comorbidities.

### Sensitivity analyses

Sensitivity analyses using alternative timeframes for severity assessment (3-year and 7-year after first AN diagnosis) identified different severe AN groups (Supplementary Table S6). We observed no significant differences between severity groups in 5- and 7-year analyses, while the severe AN group was significantly younger than the less-severe group in the 3-year severity analysis. Additionally, pharmacotherapy patterns in the severe AN groups encompassed significantly more medication categories than less-severe groups in 7-year assessments (p < 0.001), whereas the difference is not significant in the 3-year severity assessment (p = 0.484).

When AN severity was assessed 3 years post-diagnosis, the severe AN group showed lower odds than individuals with less-severe AN of receiving anti-infective drugs, drugs for the genitourinary system and sex hormones, musculoskeletal drugs, and respiratory and sensory drugs (ORs ranging from 0.79 to 0.90) but higher odds for alimentary tract medications (OR = 1.30, 95% CI 1.16-1.46), analgesics (OR = 1.14, 95% CI 1.02-1,29) and psychotropic drugs (OR = 1.63, 95% CI 1.46-1.83) (Supplementary Figure S6). The 7-year AN-RSI analysis closely resembled the primary 5-year analysis (Supplementary Figure S7). Severe AN cases had higher likelihood of being prescribed various medications such as drugs for the alimentary tract (OR = 1.70, 95% CI 1.51-1.91), cardiovascular drugs (OR = 1.27, 95% CI 1.10 −1.45), analgesics (OR = 1.20, 95% CI 1.06-1.36), and psychotropic drugs (OR = 1.97, 95% CI 1.76-2.20).

Using alternative timeframes for severity assessment also produced differences among clusters within the severe group. Although comparable clusters were identified by LCA across all cohorts (Supplementary Figures S8 and S9), the correlation between severity scores and comorbidity burden varied. Strong positive correlations were observed for the 7-year and 5-year AN-RSI (ρ = 0.99, 95% CI 0.94–1.00; ρ = 0.98, 95% CI 0.81–0.99), whereas the correlation was not statistically significant for the 3-year measure. (Supplementary Table S7).

## Discussion

AN severity is not only a key indicator for clinical outcomes but also influences pharmacological management strategies. This Danish nationwide study provides the first comprehensive characterization of pharmacotherapy patterns in severe AN. Our findings provide evidence for diverse and increased medication prescription in individuals with severe AN compared to individuals with less-severe AN, revealing–as per our hypothesis–a comorbidity-driven prescription heterogeneity within severe AN.

In our study, psychotropic medications–predominantly antidepressants, antipsychotics, anxiolytics/hypnotics, and antiepileptics–were the most frequently prescribed for severe AN. Prior research that stratified AN into “early stage” versus “severe and enduring” subtypes reported that 44.3% of AN patients were taking psychotropic medication^[19]^, though between-group differences were not explored. By contrast, an osteoporosis-focused cohort study observed an even higher psychotropic exposure rate (85.6%) in individuals with AN, with similar medication classes predominating.^[20]^ The high prescription rate is consistent with our findings and likely reflects the recruitment of a clinically severe population, mirroring our cohort’s characteristics. Among all psychotropic medications examined, significant variations were observed between severity groups. The widespread prescription of SSRI antidepressants–particularly fluoxetine and sertraline–across both severity categories likely reflects their adjunctive role in addressing comorbid mood symptoms and potential benefits for relapse prevention in weight-restored patients.^[21]^ The significantly higher olanzapine use in the severe group compared to the less-severe group corresponds with emerging evidence from large-scale randomized controlled trials demonstrating modest weight gain benefits of this medication in AN treatment.^[22]^ This may possibly reflect that individuals with severe AN present with increased risk of critically low BMI, for whom weight restoration is the highest priority. However, the relatively lower prevalence of olanzapine prescription in AN, in contrast to other psychotropic drugs, appears constrained by the absence of specific national guidelines for AN pharmacotherapy and clinician concerns regarding the metabolic side effects in young populations. This pattern also aligns with the recently updated World Federation of Societies of Biological Psychiatry (WFSBP) guidelines^[23]^, which provide only a limited recommendation for olanzapine in AN treatment, acknowledging that available evidence is restricted to weight gain effects while its impact on core AN psychopathology remains unclear. These findings reveal both notable commonality and substantial discrepancies between real-world prescribing practices and evidence-based recommendations, highlighting that clinical decisions are often influenced by multiple factors beyond guidelines.

The less frequent prescription of contraceptives in individuals with severe AN warrants clinical consideration. This prescription pattern likely reflects clinical risk stratification based on the behavioral and physiological changes associated with severe AN. Research indicates that patients with AN commonly experience sexual dysfunction, including loss of libido, increased sexual anxiety, and avoidance of sexual relationships.^[24]^ Additionally, Individuals with AN commonly experience amenorrhea due to endocrine dysfunction ^[25]^, which may give rise to reluctance to prescribe hormonal contraceptives in order to monitor the changes in the natural hormonal status of the body or reduce the perception of pregnancy risk of patients. These factors may contribute to reduced contraceptive prescribing in clinical practice. However, contrary to these findings on reduced risk of sexual activity or pregnancy, it has also been shown that women with active AN not only have an earlier age of pregnancy than the general population, but also experience higher rates of unplanned pregnancies and subsequent abortions compared to those without EDs. ^[26]^ This pattern may be explained by the lower contraceptive use observed in our study. Additionally, while contraceptives are sometimes prescribed to address bone density loss or stimulate menstruation in AN ^[27, 28]^, clinicians may hesitate to prescribe them to severe AN patients due to concerns about side effects such as increased depression risk or reduced motivation for weight restoration.^[29]^ Taken together, these findings highlight the importance of addressing contraception needs in individuals with severe AN irrespective of the presence of amenorrhea and presumed sexual activity and developing clinical guidelines that consider both reproductive health and the unique medical vulnerabilities associated with severe AN.

The diverse prescription patterns we observed in individuals with AN in the absence of comorbidities align with prior literature ^[6]^, which reports that among comorbidity-free individuals with AN, over 80% use at least one type of medication, with psychotropic drugs prescribed to more than 25% of this group. Even in the absence of a psychiatric diagnosis, clinicians may prioritize targeting subthreshold psychopathology such as pervasive anxiety, cognitive rigidity, and emotional dysregulation, all of which are frequently observed in AN and associated with poorer treatment outcomes and higher mortality.^[30]^ Notably, individuals with severe AN received disproportionally more antidepressants and antipsychotics compared to individuals with less-severe AN, reflecting the intense and diverse psychiatric symptom characteristics of severe AN: individuals with severe AN often exhibit sleep disturbances, more frequent obsessive rumination on body image, and anxiety regarding food intake. Meanwhile, a higher frequency of medications targeting the alimentary tract in severe AN also provides evidence for clinicians making considerable efforts to address the latent physiological disturbances and chronic malnutrition, as well as their focus on optimizing the outcomes for their patients with AN.

LCA provides a data-driven approach to quantify the heterogeneity of the severe AN group by identifying subgroups with distinct comorbidity profiles. The validity of clustering was supported by a similar pattern derived via K-means. Within this group, individuals with concurrent comorbidity, regardless of the level of burden, had increased use of prescriptions compared to those without comorbid diagnoses, indicating how comorbid conditions are significantly associated with prescriptions as per our hypothesis, and paralleling observations in general AN and restrictive ED populations.^[6]^ Furthermore, comparisons across clusters with comorbidity provided robust evidence for the dose-dependent association between comorbidity complexity and prescription frequency.

The timeframes used to define severity may identify distinct patient populations and influence the rationality of downstream analyses. Given the ongoing debate regarding the definition of severe AN and the novelty of applying AN-RSI in research, we conducted sensitivity analyses using alternative severity definitions: 3-year, 7-year assessments. The results of sensitivity analyses underscored the stability of our findings: the prescription patterns of the severe AN group were illustrated as similarly directional patterns when different assessment timepoints were used, even though the consistently attenuated effect sizes were found from the 3-year severity analyses. Moreover, the validity of our severity classification at 5 years post first AN diagnosis was supported by multiple findings, which were detailed in the Supplementary Materials.

To the best of our knowledge, this is the first study to systematically investigate pharmacotherapy practices in severe AN with a primary focus on identifying distinct prescription patterns associated with illness severity. Our work has several additional strengths. First, the use of national health registry data enabled us to capture prescription records across an unselected population of individuals with AN, minimizing selection bias. Second, the longitudinal design with up to 10 years of follow-up allowed us to analyze trends in prescribing practices over time. Third, stratifying patients into subgroups based on comorbidity profiles that incorporated not only psychiatric but also somatic conditions provided novel insights into heterogeneity in medication prescription patterns, acknowledging that complications arising later in the illness course may also be associated with the complexity of prescription patterns.

While this study advances our understanding of real-world pharmacotherapy in severe AN, key limitations exist. First, differences in dispensed prescriptions between groups may be underestimated since prescriptions during inpatient admissions or specialist care are not captured in the prescription register (linked to CPR). Given higher hospitalization rates in severe AN, this could result in underreporting of hospital-related prescriptions. Second, over-the-counter drugs (e.g., gastrointestinal agents, nutritional supplements) were not included, potentially underrepresenting adjunctive treatments. Third, clinical data such as symptoms, body mass index, or laboratory parameters were unavailable, limiting our ability to adjust for potential confounders; for example, malnutrition or a lower body mass index may drive supplement prescriptions, thereby confounding the associations between severity and prescription. Fourth, the clinical indication for each prescription was unknown, and bidirectional associations between AN and both somatic and psychiatric comorbidities further complicated the interpretation of prescribing patterns. On the one hand, psychotropic medications were frequently prescribed despite not being specifically recommended for the treatment of anorexia nervosa, and unknown indications prevent clear interpretation of whether prescribing reflects treatment of comorbid conditions or potential guideline non-adherence. On the other hand, somatic diagnoses were not differentiated by clinical chronicity or treatment intent (e.g., transient diagnoses versus chronic conditions related to AN that require ongoing treatment). This reflects a deliberate design choice to capture overall medication burden but limits the clinical interpretability of specific somatic drug classes. Finally, the generalizability of this study may be limited due to differences in clinical practices across healthcare systems; validation in other countries is warranted.

In conclusion, by mapping real-world medication prescriptions in a national cohort, our study reveals the complex relationship between AN severity, the presence of multiple comorbidities, and medication prescription patterns. We anticipate our findings will bring further clarity to the complexity of managing AN and the necessity of evidence-based treatment guidelines, especially for severe AN. We also urge both clinicians and researchers to pay greater attention to the substantial heterogeneity in comorbidity profiles among individuals with severe AN and to develop comprehensive and specialized treatment strategies for this vulnerable population.

## ACKNOWLEDGMENTS

Zeynep Yilmaz served as Principal Investigator and acts as the guarantor of the work. She accepts full responsibility for the integrity of the data, the accuracy of the analyses, and the decision to publish. Study concept and design: All authors. Acquisition, analysis, or interpretation of data: ZZ, HC, JTL, and ZY. Drafting of the manuscript: ZZ and ZY. Critical revision of the manuscript for important intellectual content: All authors. Statistical analysis: ZZ, HC, JTL.

## COMPETING INTERESTS

The authors have declared no competing interests.

## FUNDING STATEMENT

This study was funded by the Danmarks Frie Forskningsfond (Independent Research Fund Denmark; Sapere Aude awarded to Zeynep Yilmaz, case no. 1052-00029B) and by the U.S. Department of Health and Human Services, National Institutes of Health, National Institute of Mental Health (grant R01MH136156, Eating Disorders Genetics Initiative 2).

## DATA AVAILABILITY

Data for this study were obtained from Danish national registries via Statistics Denmark and the Danish Health Data Authority. Individual-level data cannot be shared due to Danish data protection legislation and registry access agreements. Qualified researchers may apply for data access through the appropriate Danish authorities, subject to approval of research protocols and compliance with data protection requirements. Statistical analysis code is available from the corresponding author upon request. This work was previously made available as a preprint on medRxiv (doi: 10.1101/2025.07.15.25330554).

## ETHICS APPROVAL

This study was approved by the Danish Data Protection Agency (“AU fællesanmeldelse” journal number 2016-051-000001 serial number 2277) and the Danish Health Data Authority. According to Danish law, review by an ethics board and patient consent are not required for purely register-based studies. All data were de-identified and not recognizable at an individual level.

## Notes

### Funding Statement

This study was funded by the Independent Research Fund Denmark (DFF) Sapere Aude awarded to ZY (case no. 1052-00029B) and the US National Institute of Mental Health (NIMH) Eating Disorders Genetics Initiative 2 (R01MH136156).
ZY acknowledges grant support from the DFF (case no. 1052-00029B, 3166-00063B, and 4309-00050B), Lundbeck Foundation Ascending Investigator (R434-2023-269), and NIMH (R01MH136156). LVP acknowledges Novo Nordisk Fonden, Grant/Award Number: NNF20OC0064993. BJV is supported by Lundbeck Foundation (R335-2019-2339), and the Independent Research Fund Denmark (2034-00241B).

### Author Declarations

This study was approved by the Danish Data Protection Agency, and the Danish Health Data Authority. Each scientific project must be approved before initiation, and approval is granted to a specific Danish research institution. According to Danish law, review by an ethics board and patient consent are not required for purely register-based studies. All data were de-identified and not recognizable at an individual level.

### Summary of Updates

This version of the manuscript has been revised at Jan 12th 2026.

## REFERENCES

1. van Eeden, A.E., D. van Hoeken, and H.W. Hoek, Incidence, prevalence and mortality of anorexia nervosa and bulimia nervosa. Curr Opin Psychiatry, 2021. 34(6): p. 515–524.

2. Herpertz-Dahlmann, B., et al., Outcome of childhood anorexia nervosa-The results of a five- to ten-year follow-up study. Int J Eat Disord, 2018. 51(4): p. 295–304.

3. Dobrescu, S.R., et al., Anorexia nervosa: 30-year outcome. Br J Psychiatry, 2020. 216(2): p. 97–104.

4. Touyz, S., et al., Treating severe and enduring anorexia nervosa: a randomized controlled trial. Psychol Med, 2013. 43(12): p. 2501–11.

5. Crow, S.J., Pharmacologic Treatment of Eating Disorders. Psychiatr Clin North Am, 2019. 42(2): p. 253–262.

6. Clausen, L., et al., Pharmacotherapy in anorexia nervosa: A Danish nation-wide register-based study. J Psychosom Res, 2023. 164: p. 111077.

7. Udo, T. and C.M. Grilo, Psychiatric and medical correlates of DSM-5 eating disorders in a nationally representative sample of adults in the United States. Int J Eat Disord, 2019. 52(1): p. 42–50.

8. Westmoreland, P., M.J. Krantz, and P.S. Mehler, Medical Complications of Anorexia Nervosa and Bulimia. Am J Med, 2016. 129(1): p. 30–7.

9. Larsen, J.T., et al., Construction of a register-based severity index for anorexia nervosa in Denmark: Association with overall and cause-specific mortality. Eur Psychiatry, 2025. 68(1): p. e104.

10. Broomfield, C., et al., Labeling and defining severe and enduring anorexia nervosa: A systematic review and critical analysis. Int J Eat Disord, 2017. 50(6): p. 611–623.

11. Hay, P. and S. Touyz, Classification challenges in the field of eating disorders: can severe and enduring anorexia nervosa be better defined? J Eat Disord, 2018. 6: p. 41.

12. Lynge, E., J.L. Sandegaard, and M. Rebolj, The Danish National Patient Register. Scand J Public Health, 2011. 39(7 Suppl): p. 30–3.

13. Mors, O., G.P. Perto, and P.B. Mortensen, The Danish Psychiatric Central Research Register. Scand J Public Health, 2011. 39(7 Suppl): p. 54–7.

14. Wildes, J.E., et al., Characterizing severe and enduring anorexia nervosa: An empirical approach. Int J Eat Disord, 2017. 50(4): p. 389–397.

15. Dalle Grave, R., Severe and enduring anorexia nervosa: No easy solutions. Int J Eat Disord, 2020. 53(8): p. 1320–1321.

16. Kildemoes, H.W., H.T. Sorensen, and J. Hallas, The Danish National Prescription Registry. Scand J Public Health, 2011. 39(7 Suppl): p. 38–41.

17. Pedersen, C.B., The Danish Civil Registration System. Scand J Public Health, 2011. 39(7 Suppl): p. 22–5.

18. WHO Collaborating Centre for Drug Statistics Methodology, Guidelines for ATC classification and DDD assignment,. 2025; Available from: https://atcddd.fhi.no/atc_ddd_index_and_guidelines/guidelines/.

19. Ambwani, S., et al., A multicenter audit of outpatient care for adult anorexia nervosa: Symptom trajectory, service use, and evidence in support of “early stage” versus “severe and enduring” classification. Int J Eat Disord, 2020. 53(8): p. 1337–1348.

20. Garner, D.M., et al., Psychotropic medications in adult and adolescent eating disorders: clinical practice versus evidence-based recommendations. Eat Weight Disord, 2016. 21(3): p. 395–402.

21. Marvanova, M. and K. Gramith, Role of antidepressants in the treatment of adults with anorexia nervosa. Ment Health Clin, 2018. 8(3): p. 127–137.

22. Attia, E., et al., Olanzapine Versus Placebo in Adult Outpatients With Anorexia Nervosa: A Randomized Clinical Trial. Am J Psychiatry, 2019. 176(6): p. 449–456.

23. Himmerich, H., et al., World Federation of Societies of Biological Psychiatry (WFSBP) guidelines update 2023 on the pharmacological treatment of eating disorders. World J Biol Psychiatry, 2023. 24(8): p. 643–706.

24. Price, T., et al., Sexual function and dysfunction among women with anorexia nervosa: A systematic scoping review. Int J Eat Disord, 2020. 53(9): p. 1377–1399.

25. Golden, N.H. and I.R. Shenker, Amenorrhea in anorexia nervosa. Neuroendocrine control of hypothalamic dysfunction. Int J Eat Disord, 1994. 16(1): p. 53–60.

26. Bulik, C.M., et al., Unplanned pregnancy in women with anorexia nervosa. Obstet Gynecol, 2010. 116(5): p. 1136–40.

27. Maimoun, L., et al., Oral contraceptives partially protect from bone loss in young women with anorexia nervosa. Fertil Steril, 2019. 111(5): p. 1020–1029 e2.

28. Golden, N.H., et al., Resumption of menses in anorexia nervosa. Arch Pediatr Adolesc Med, 1997. 151(1): p. 16–21.

29. Skovlund, C.W., et al., Association of Hormonal Contraception With Depression. JAMA Psychiatry, 2016. 73(11): p. 1154–1162.

30. Fewell, L.K., C.A. Levinson, and L. Stark, Depression, worry, and psychosocial functioning predict eating disorder treatment outcomes in a residential and partial hospitalization setting. Eat Weight Disord, 2017. 22(2): p. 291–301.

