## Supplementary materials for "Medication Use in Severe Anorexia Nervosa: A Danish Register-based Study"

Content


### Sensitivity analyses

Discussion

### Tables

S1. Anorexia Nervosa Register-based Severity Index

S2. Medication classifications and Anatomical Therapeutic Chemical (ATC) codes

S3. Definition of psychiatric and somatic comorbidity

S4. Model fit statistics for latent class analysis

S5. Characteristics of patients with severe and non-severe anorexia nervosa

S6. Characteristics of patients with severe vs less-severe anorexia nervosa (severity evaluated at 3- or 7-year post-diagnosis)

S7. Association between comorbidity clusters and severity scores at different assessment time points

### Figures

S1. Figure illustration of study design

S2. Analgesics and psychotropic drugs in severe vs less-severe anorexia nervosa patients

S3. Medication prescriptions in severe vs less-severe anorexia nervosa patients without comorbidities

S4. Comorbidity profiles of patients with severe anorexia nervosa

S5. Medication prescriptions in severe anorexia nervosa patients with distinct comorbidity profiles

S6. Medication prescriptions in patients with severe vs less-severe anorexia nervosa between 5 and 10 years post-diagnosis (severity evaluated at 3 years post-diagnosis)

S7. Medication prescriptions in patients with severe vs less-severe anorexia nervosa between 5 and 10 years post-diagnosis (severity evaluated at 7 years post-diagnosis)

S8. Comorbidity profiles of patients with severe anorexia nervosa (severity evaluated at 3 years post-diagnosis, derived from latent class analysis)

S9. Comorbidity profiles of patients with severe anorexia nervosa (severity evaluated at 7 years post-diagnosis, derived from latent class analysis)

### Discussion

Sensitivity analyses not only provided evidence for the robustness of our findings on prescription patterns of the severe anorexia nervosa (AN) group, but also indicate the validity of severity classification with the Anorexia Nervosa Register-based Severity Index (AN-RSI) assessed at 5 years post first AN diagnosis. Several key findings support the need for a longer assessment period. First, compared to the 5- and 7-year assessments, the 3-year assessment failed to distinguish between severe and less-severe groups in terms of total medication categories, indicating that a shorter assessment period may not be sufficient to capture the full picture of complex prescription patterns in severe AN. Second, age of onset patterns differed between timeframes: the 3-year assessment identified younger patients having higher severity scores, while other timeframes showed similar age distribution among severity groups. This finding suggests that the 3-year assessment may not represent the complexity of late-onset presentations, given that the original AN-RSI accounts for both early and late onset as severity indicators with equal weighting. Third, when the 3-year and 7-year periods were used to assess severity, while findings from both analyses showed similar directional patterns compared to the primary 5-year analysis, the consistently attenuated effect sizes from the 3-year severity analysis suggested less distinction between severity groups using a shorter timeframe. Fourth, in contrast to the consistent correlations between severity and comorbidity burden found in the 5- or 7-year defined severe AN group, the 3-year assessment generates a heterogeneous “severe” group that includes patients who may not truly be severe. This difference likely reflects the insufficient time to capture overall severity and late-emerging complications during illness progression. The analyses with 5-year and 7-year severity assessments, which mirror thresholds commonly used in longitudinal studies, showed consistent findings, demonstrating their ability to better identify cohesive severe populations.

Taken together, our study underscores the benefit of extended timeframes (5-7 years) for severity classifications with AN-RSI, which better characterize severe AN groups and reflect cumulative illness burden more accurately than 3 years and therefore reduce misclassification in samples including adolescent and adult patients. We specifically recommend 5 years to balance the need for precise severity assessment with avoiding excessively long observation periods, which might delay the identification of severe cases requiring early intervention.

It should be noted that our recommendations are based on findings from the present study, with a specific focus on the medication prescription pattern and a sample spanning a range of ages. Further studies are needed to confirm that the 5-year severity assessment could similarly capture the unique characteristics of severe AN in terms of genetic architecture, risk factor profiles, and long-term outcomes across different age groups.

Supplementary Table S1.

### Anorexia Nervosa Register-based Severity Index

| Variable | Definition | Weight |
|---|---|---|
| Early onset | First AN-related hospital contact (inpatient or outpatient) before age 10. | 1 point. |
| Late onset | First AN-related hospital contact (inpatient or outpatient) after age 25. | 1 point. |
| Inpatient admissions | Count of recorded inpatient hospital admissions with AN diagnosis. | 2 points per admission. |
| Outpatient contacts | Number of outpatient treatment contacts with an AN diagnosis, excluding initial treatment contact. | 1 point per contact. |
| Treatment length | Length of aggregate period of inpatient and outpatient treatment.<br><br>For inpatient admission: calculated as interval between admission and discharge dates.<br><br>For outpatient episodes: calculated as 0.5 × (interval between start and end date) + 0.5 × total treatment days | 1 point per complete year. |
| Illness duration | Time from initial AN diagnosis to most recent treatment day within the 5-year follow-up period (primary definition) or until each specific treatment timepoint (continuously updated definition used in sensitivity analyses). | 1 point per complete year. |

*Note*: AN: anorexia nervosa

Supplementary Table S2a.

### Medication classifications and Anatomical Therapeutic Chemical (ATC) Codes

| Medication Class | Medication Subclass | ATC Code |
|---|---|---|
| Alimentary tract (A) | Gastrointestinal | A02-A07 |
| | Vitamins/Minerals | A11/A12 |
| Cardiovascular drugs (B, C) | Iron preparations | B03A |
| Anti-infective drugs (D,J,P) | | |
| Genitourinary System and Sex Hormones (G) | Sex Hormones preparations | G03 |
| | Hormonal contraceptives | G03A |
| Hormonal preparations (H)[a] | Thyroid therapy | H03 |
| Immunomodulating drugs (I) | | |
| Musculoskeletal drugs (M) | Bisphosphonates | M05 |
| | NSAIDs | M01A |
| Analgesics (N02) | Analgesics | N02 |
| Psychotropic drugs (N, except for N02) | Antiepileptics | N03 |
| | Antipsychotics | N05A |
| | Typical antipsychotics | N05AA-N05AD, N05AF, N05AG |
| | Atypical antipsychotics | N05AH, N05AL, N05AX |
| | Anxiolytics/Hypnotics | N05B/N05C |
| | Antidepressants | N06A |
| | SSRI | N06AB |
| | non-SSRI | N06AA, N06AF, N06AG, N06AX |
| | Psychostimulants | N06B |
| | Other NS drugs | N01, N04, N07 |
| Respiratory and sensory drugs (R,S) | | |
| Other (V) | | |

*Note:* Medication subclasses of non-psychotropic drugs were selected at the first three to four digits of the ATC code to ensure adequate sample size and statistical power, and were chosen based on the high prevalence of relevant comorbidities (e.g., bisphosphonates for osteoporosis, iron preparations for anemia). NSAIDs: nonsteroidal anti-inflammatory drugs; SSRI: selective serotonin reuptake inhibitors. [a] Hormonal preparations category (H) excludes sex hormones and insulin.

Supplementary Table S2b.

### Psychotropic drugs and Anatomical Therapeutic Chemical (ATC) codes

| Medication Class | Medication | ATC Code |
|---|---|---|
| Analgesics | Codeine combinations[a] | N02AA59 |
| | Tramadol | N02AX02 |
| | Paracetamol | N02BE01 |
| Antiepileptics | Lamotrigine | N03AX09 |
| Antipsychotics (Typical) | Chlorprothixene | N05AF03 |
| Antipsychotics (Atypical) | Olanzapine | N05AH03 |
| | Quetiapine | N05AH04 |
| | Risperidone | N05AX08 |
| Anxiolytics/Hypnotics | Diazepam | N05BA01 |
| | Oxazepam | N05BA04 |
| | Zopiclone | N05CF01 |
| | Melatonin | N05CH01 |
| Antidepressants (SSRI) | Fluoxetine | N06AB03 |
| | Citalopram | N06AB04 |
| | Paroxetine | N06AB05 |
| | Sertraline | N06AB06 |
| | Escitalopram | N06AB10 |
| Antidepressants (non-SSRI) | Mirtazapine | N06AX11 |
| | Venlafaxine | N06AX16 |
| Psychostimulants | Methylphenidate | N06BA04 |

*Note:* Medications included in the table represent the 20 most frequently prescribed drugs from the N chapter of ATC in the cohort, selected to allow meaningful comparisons between severe and less-severe AN groups. SSRI: Selective Serotonin Reuptake Inhibitor; [a] Codeine combination, excl. antipsychotics (N05)

Supplementary Table S3a.

### Definition of psychiatric comorbidity

| **Comorbidity** | **International Classification of Disease (ICD)** ICD-10 | ICD-8 | **Definition** |
|---|---|---|---|
| Psychiatric disorders: DSM-IV Axis I | | | |
| Substance Use | F10-F19 | 291.x9, 294.39, 303.x9, 303.20, 303.28, 303.90, 304.x9 | Having diagnosis during the period from first AN diagnosis to 10 years afterwards. |
| Mood | F30-F39 | 296.x9 (excluding 296.89), 298.09, 298.19, 300.49, 301.19 | |
| Anxiety | F40-F48 | 300.x9 (excluding 300.49), 305.x9, 305.68, 307.99 | |
| Other psychiatric disorders | | | |
| Organic | F00-F09 | 290.09, 290.10, 290.11, 290.18, 290.19, 292.x9, 293.x9, 294.x9, 309.x9 | Having diagnosis during the period from age 6 to 10 years after first AN diagnosis. |
| Schizophrenia | F20-F29 | 295.x9, 296.89, 297.x9, 298.29-298.99, 299.04, 299.05, 299.09, 301.83 | |
| Sleep | F51 | 306.49 | |
| Personality | F60-F62 | 301.x9, 301.80-301.84 | |
| Habit impulse | F63 | | |
| Intellectual | F70-F79 | 311-315 | |
| Developmental | F84 | 299.00, 299.01, 299.02, 299.03 | |
| Behavioral and emotional | F90-F98 | 306·x9, 308·0x | |
| Self-harm | T36-T65, S50, S51, S55, S59, S60, S61, S65, S69, X60-X84 | E950-959 | Having diagnosis during the period from age 6 to 10 years after first AN diagnosis. |

*Note:* Rationale for Psychiatric Comorbidity Assessment: The distinction between DSM-IV Axis I disorders and other psychiatric disorders acknowledges the differential impact these conditions may have on AN treatment approaches. Mood, anxiety, and substance use disorders frequently co-occur with AN and may directly influence both disease progression and medication selection. Axis I disorders were captured longitudinally from the first AN diagnosis until ten years thereafter, allowing inclusion of comorbidities that may develop in response to or concurrently with AN. Other psychiatric disorders, including personality and intellectual/developmental disorders, were captured from age six, reflecting their potential developmental origins and pre-existing influence on treatment decisions.

Supplementary Table S3b.

### Definition of somatic comorbidity

| Comorbidity | International Classification of Diseases (ICD) ICD-10 | ICD-8 | Definition |
|---|---|---|---|
| Somatic diseases | | | |
| Respiratory | J00-J46, R05-R06 | 460-466, 470-486, 490-493, 502-508, 783 | Having diagnosis during the period from age 6 to 10 years after first AN diagnosis. |
| Infection | A00-B99 | 000-136 | |
| Musculoskeletal | M00-M68 | 710-718 | |
| Gastronintestinal | K25-K37,K70-K85 | 530-535, 540-542, 563, 570-577 | |
| Endocrine | E00-E35 | 240-246,250-259 | |
| Skin | L00-L54, L90-L95 | 680-690,701,708-709 | |
| Neurological | G00-G47, G60-G73, G91 | 320-358 | |
| Circulatory | I00-I79, I98.3 | 390-448, 450 | |
| Congenital malformations | Q00-Q99 | 740-759 | |
| Genitourinary | N00-N08, N10-N12, N20-N21, N41, N45 | 580-584, 590, 592, 594, 601, 604 | |
| Immune | D80-D89 | | |
| Injuries | S00-T35, T66-T98, V01-V99, X85-Y98 | N800-N999, E807-E949, E960-E999 | |

*Note:* Rationale for Somatic Comorbidity Assessment: Recording somatic conditions from age six captures pre-existing medical conditions that may complicate AN treatment or influence medication selection. The extended follow-up (until ten years post-diagnosis) allows for identification of emerging somatic complications that may emerge as a consequence of AN and that could necessitate additional pharmacological interventions. This comprehensive timeframe acknowledges that somatic comorbidities may be both contributors to and consequences of severe AN, each potentially affecting the medication prescribing patterns.

Supplementary Table S4.

### Model fit statistics for latent class analysis

| N | AIC | BIC | Maximum Likelihood | Entropy | Sample Proportion |
|---|---|---|---|---|---|
| 2 | 24706.7 | 24966.5 | -12304.4 | 8.3057 | 0.23/0.77 |
| 3 | 24587.7 | 24980.0 | -12219.8 | 8.2645 | 0.33/0.21/0.46 |
| 4 | 24526.6 | 25071.5 | -12174.3 | 8.2173 | 0.47/0.11/0.31/0.11 |
| 5 | 24542.3 | 25199.7 | -12147.1 | 8.1913 | 0.03/0.05/0.04/0.19/0.7 |

*Note:* AIC = Akaike information criterion; BIC = Bayesian Information Criterion. The 4-class solution was selected based on an optimal balance between statistical fit indices, clinical interpretability, and to ensure each cluster contained more than 10% of the total sample.

Supplementary Table S5.

### Characteristics of patients with severe and non-severe anorexia nervosa

| Characteristics | Severe AN (%)[1] | Less-severe AN (%)[1] | P-value[2] |
|---|---|---|---|
| N | 1652 (21.6) | 6002 (78.4) | |
| Sex | | | 0.004 |
| Female | 1572 (95.2) | 5592 (93.2) | |
| Male | 80 (4.8) | 410 (6.8) | |
| Age at AN diagnosis (years) | 19.6 (6.7) | 19.7 (6.2) | 0.481 |
| Standardized AN severity score | 1.40 (1.4) | -0.39 (0.3) | <0.001 |
| Had any comorbidity | 1483 (89.8) | 4880 (81.3) | <0.001 |
| Had any psychiatric comorbidity | 1175 (71.1) | 3219 (53.6) | <0.001 |
| Organic disorders | 35 (2.1) | 66 (1.1) | 0.002 |
| Substance use disorders | 199 (12.1) | 529 (8.8) | <0.001 |
| Schizophrenia | 231 (14.0) | 408 (6.8) | <0.001 |
| Mood disorders | 594 (36.0) | 1399 (23.3) | <0.001 |
| Anxiety disorders | 641 (38.8) | 1667 (27.8) | <0.001 |
| Sleep disorders | 9 (0.5) | 18 (0.3) | 0.137 |
| Personality disorders | 575 (34.8) | 1350 (22.5) | <0.001 |
| Intellectual disorders | 38 (2.3) | 89 (1.5) | 0.028 |
| Developmental disorders | 87 (5.3) | 173 (2.9) | <0.001 |
| Behavioral disorders | 231 (14.0) | 549 (9.1) | <0.001 |
| Habit impulse | 10 (0.6) | 10 (0.2) | 0.002 |
| Had any somatic comorbidity | 1224 (74.1) | 3989 (66.5) | <0.001 |
| Infectious | 224 (13.6) | 697 (11.6) | 0.035 |
| Endocrine | 176 (10.7) | 400 (6.7) | <0.001 |
| Injuries | 604 (36.6) | 2132 (35.5) | <0.452 |
| Neurological | 115 (7.0) | 354 (5.9) | 0.124 |
| Circulatory | 135 (8.2) | 273 (4.5) | <0.001 |
| Respiratory | 281 (17.0) | 951 (15.8) | 0.027 |
| Gastrointestinal | 158 (9.6) | 460 (7.7) | <0.001 |
| Skin | 201 (12.2) | 445 (7.4) | <0.001 |
| Musculoskeletal | 302 (18.3) | 1223 (20.4) | 0.064 |
| Genitourinary | 50 (3.0) | 158 (2.6) | 0.431 |
| Congenital | 104 (6.3) | 310 (5.2) | 0.082 |
| Self-harm | 451 (27.3) | 933 (15.5) | <0.001 |
| Any medication | 1504 (99) | 5958 (98) | 0.017 |
| Total number of medications | 4.2 (1.7) | 3.8 (1.7) | <0.001 |
| Calendar year | | | 0.010 |
| Before 1994 | 47 (23.5) | 153 (76.5) | |
| 1994-2005 | 722 (20.1) | 2876 (79.9) | |
| After 2005 | 883 (21.6) | 2973 (77.1) | |

AN: anorexia nervosa

[1]n (%); Mean (SD); Median (25th-75th percentile)

[2]Pearson's Chi-squared test; Wilcoxon rank sum test; Fisher's exact test

Supplementary Table S6.

### Characteristics of patients with severe vs less-severe anorexia nervosa (severity evaluated at 3- or 7-year post-diagnosis)

| Severity Assesment | Characteristics | Severe AN (%)[1] | Less-severe AN (%)[1] | P-value[2] |
|---|---|---|---|---|
| 3-year post-diagnosis | N | 1655 (21.9) | 5913 (78.1) | |
| | Sex | | | 0.155 |
| | Female | 1563 (94.4) | 5525 (93.4) | |
| | Male | 92 (5.6) | 388 (6.6) | |
| | Age at AN diagnosis (years) | 19.15 (6.54) | 19.88 (6.28) | <0.001 |
| | Standardized AN severity score | 1.37 (1.36) | -0.38 (0.30) | <0.001 |
| | Total number of medications | 4.1 (1.8) | 4.1 (1.7) | 0.484 |
| 7-year post-diagnosis | N | 1719 (22.2) | 6014 (77.8) | |
| | Sex | | | <0.001 |
| | Female | 1642 (95.5) | 5596 (93.0) | |
| | Male | 77 (4.5) | 418 (7.0) | |
| | Age at AN diagnosis (years) | 19.8 (6.7) | 19.6 (6.1) | 0.203 |
| | Standardized AN severity score | 1.3 (1.4) | -0.4 (0.3) | <0.001 |
| | Total number of medications | 3.7 (2.0) | 3.4 (1.9) | <0.001 |

*Note:* Sample sizes varied due to data availability in the prescription register. The cohort for the 3-year assessment included individuals with an AN diagnosis recorded between 1993 and 2011, with follow-up ending on 31 December 2021. The cohort for the 7-year assessment included individuals with an AN diagnosis recorded between 1989 and 2011, with follow-up also ending on 31 December 2021. AN: anorexia nervosa. [1]n (%); Mean (SD); Median (25th-75th percentile) [2]Pearson's Chi-squared test; Wilcoxon rank sum test; Fisher's exact test

Supplementary Table S7.

### Association between comorbidity clusters and severity scores at different assessment time points

| Comorbidity Clusters | AN-RSI Scores[1] by Years Post-diagnosis | | |
|---|---|---|---|
| | 3 Years | 5 Years | 7 Years |
| Cluster 1[a] | 1.13 (1.15) | 1.18 (1.17) | 1.13 (1.15) |
| Cluster 2[b] | 1.15 (1.25) | 1.22(1.30) | 1.21 (1.45) |
| Cluster 3[c] | 1.29 (1.07) | 1.36 (1.27) | 1.31(1.39) |
| Cluster 4[d] | 1.64 (0.95) | 1.64 (1.42) | 1.50 (1.34) |
| Cluster 5[e] | 1.54 (1.27) | 1.81 (1.31) | 1.86 (1.64) |
| Correlation Coefficient (95% CI)[2] | 0.85(-0.1-0.99) | 0.98(0.81-0.99) | 0.99(0.94-1.00) |
| Correlation Coefficient P-value | 0.069 | 0.001 | <0.001 |

*Note.* AN-RSI: Anorexia Nervosa Register-based Severity Index
[1] Mean (standard deviation);
[2] Clusters were treated as ordinal categorical variables. Spearman's rank correlation coefficients between comorbidity clusters and AN-RSI scores are reported in the bottom row.
[a] Cluster 1: Severe AN patients without any comorbidities
[b] Cluster 2: Severe AN patients with low burden of comorbidities
[c] Cluster 3: Severe AN patients with medium burden of personality, mood, and anxiety disorders
[d] Cluster 4: Severe AN patients with high burden of personality disorders and self-harm conditions
[e] Cluster 5: Severe AN patients with high burden comorbidities

Supplementary Figure S1.

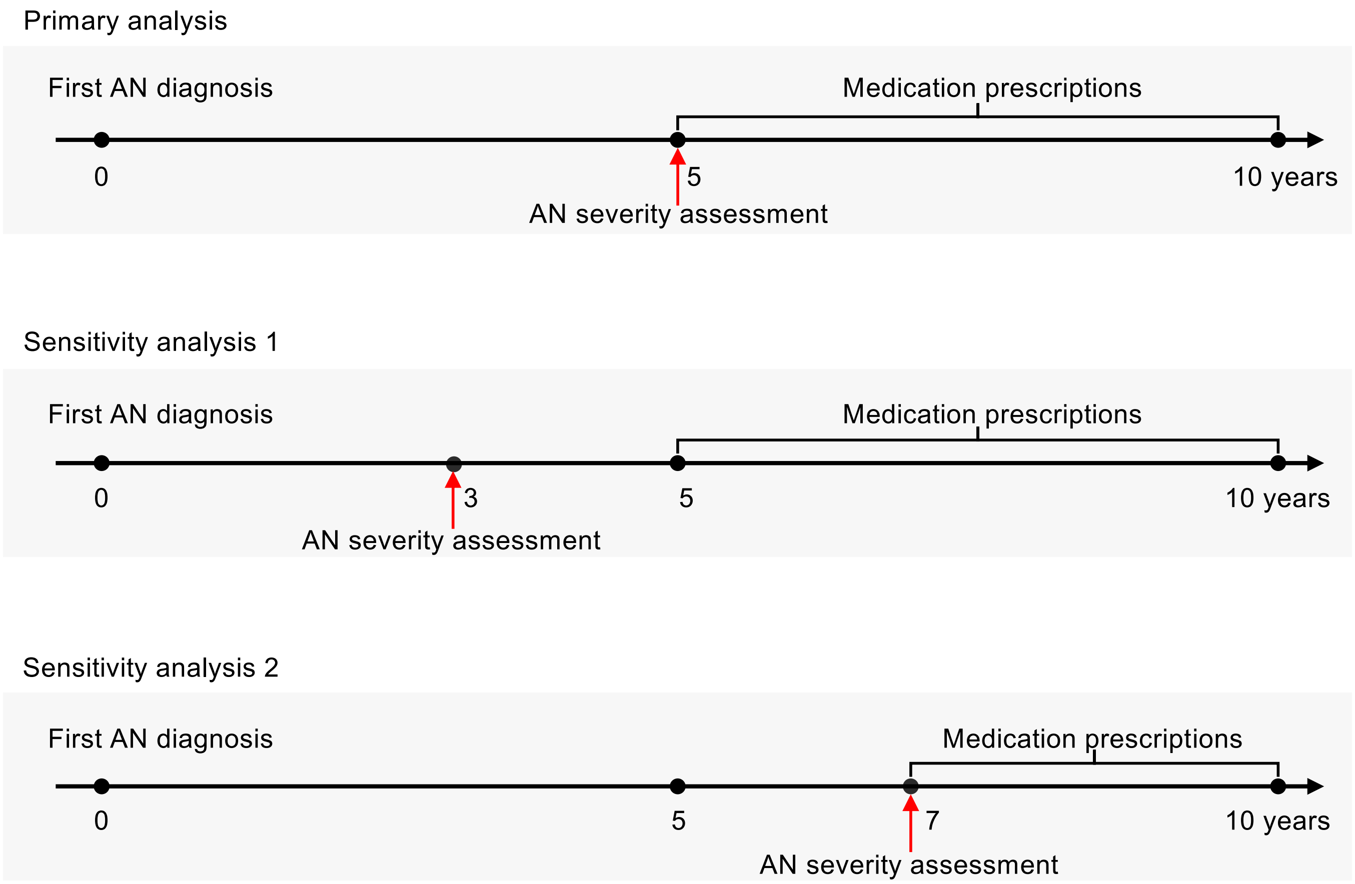

Supplementary Figure S2.

### Analgesics and psychotropic drugs in severe vs less-severe anorexia nervosa patients

| Medication | Severe AN (%) | Less–severe AN (%) | Odds Ratio (95% CI) | P–value |
|---|---|---|---|---|
| **Analgesics** | | | | |
| Codeine combinations | 2.7 | 2.3 | 1.17(0.83–1.65) | 0.142 |
| Tramadol | 10.3 | 9.0 | 1.16(0.97–1.39) | 0.056 |
| Paracetamol | 16.1 | 12.8 | 1.31(1.13–1.53) | < 0.001 |
| **Antiepileptics** | | | | |
| Lamotrigine | 6.2 | 3.4 | 1.87(1.47–2.39) | < 0.001 |
| **Typical antipsychotics** | | | | |
| Chlorprothixene | 10.7 | 4.1 | 2.79(2.28–3.41) | < 0.001 |
| **Atypical antipsychotics** | | | | |
| Olanzapine | 7.9 | 2.2 | 3.74(2.92–4.79) | < 0.001 |
| Risperidone | 5.2 | 2.2 | 2.38(1.80–3.13) | < 0.001 |
| Quetiapine | 13.1 | 6.1 | 2.34(1.96–2.80) | < 0.001 |
| **SSRI Antidepressants** | | | | |
| Sertraline | 20.7 | 10.6 | 2.21(1.91–2.55) | < 0.001 |
| Fluoxetine | 12.3 | 6.1 | 2.17(1.81–2.60) | < 0.001 |
| Citalopram | 11.6 | 10.3 | 1.15(0.96–1.36) | 0.066 |
| Paroxetine | 2.7 | 1.9 | 1.45(1.02–2.07) | 0.024 |
| Escitalopram | 5.0 | 4.5 | 1.12(0.87–1.44) | 0.145 |
| **Non–SSRI Antidepressants** | | | | |
| Mirtazapine | 9.1 | 5.8 | 1.63(1.33–1.99) | < 0.001 |
| Venlafaxine | 9.3 | 5.3 | 1.84(1.51–2.25) | < 0.001 |
| **Anxiolytics/Hypnotics** | | | | |
| Oxazepam | 12.2 | 5.5 | 2.38(1.98–2.86) | < 0.001 |
| Diazepam | 2.7 | 2.2 | 1.23(0.87–1.73) | 0.010 |
| Zopiclone | 10.2 | 6.6 | 1.60(1.32–1.93) | < 0.001 |
| Melatonin | 7.4 | 3.4 | 2.24(1.78–2.83) | < 0.001 |
| **Psychostimulants** | | | | |
| Methylphenidate | 2.9 | 3.0 | 0.96(0.70–1.34) | 0.551 |

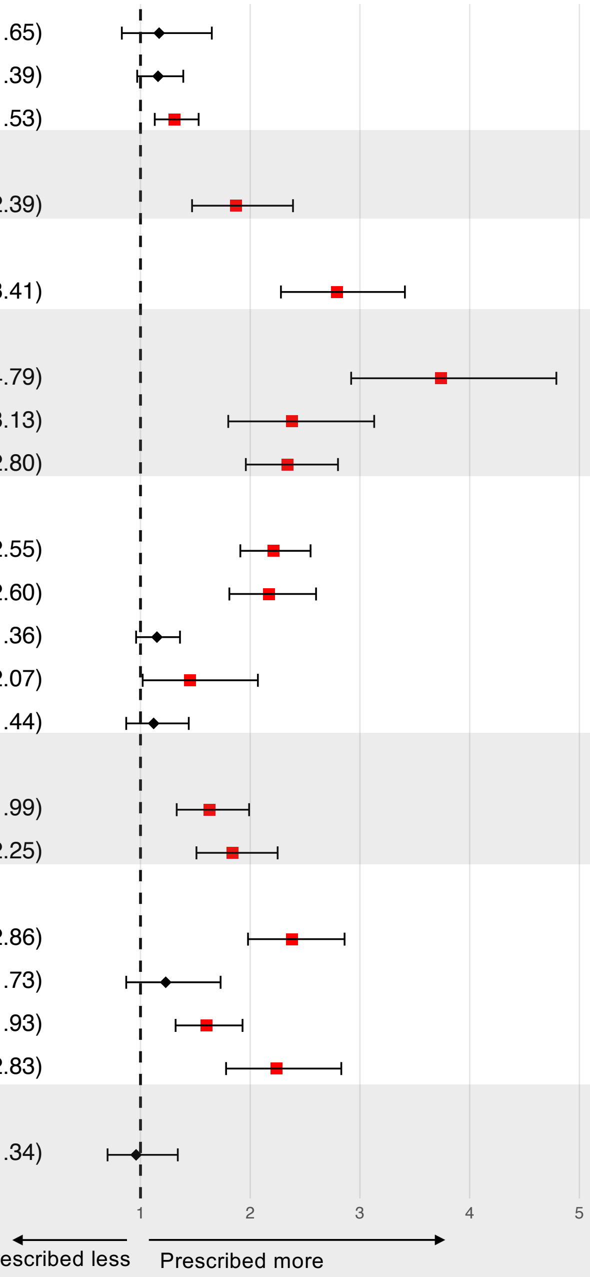

Supplementary Figure S3.

### Medication prescriptions in severe vs less-severe anorexia nervosa patients without comorbidities

| Medication | Severe AN (%) | Less–severe AN (%) | Odds Ratio (95% CI) | P–value |
|---|---|---|---|---|
| Alimentary tract | 25.4 | 19.5 | 1.41(0.97–2.05) | 0.046 |
| Gastrointestinal | 20.7 | 15.7 | 1.40(0.94–2.11) | 0.062 |
| Vitamins/Minerals | 5.9 | 2.2 | 2.76(1.30–5.85) | 0.006 |
| Cardiovascular drugs | 11.8 | 13.5 | 0.86(0.52–1.42) | 0.323 |
| Anti–infective drugs | 85.2 | 87.6 | 0.81(0.51–1.29) | 0.226 |
| Genitourinary system and sex hormones | 65.1 | 76.0 | 0.59(0.42–0.83) | 0.002 |
| Sex hormones | 63.3 | 69.7 | 0.75(0.54–1.05) | 0.057 |
| Hormonal preparation drugs | 3.0 | 3.9 | 0.75(0.29–1.91) | 0.346 |
| Musculoskeletal drugs | 19.5 | 22.8 | 0.82(0.55–1.23) | 0.196 |
| NSAID | 19.5 | 22.6 | 0.83(0.55–1.24) | 0.21 |
| Analgesics | 13.0 | 11.1 | 1.20(0.74–1.96) | 0.267 |
| Psychotropic drugs | 47.3 | 27.3 | 2.40(1.72–3.33) | < 0.001 |
| Antipsychotics | 5.3 | 2.0 | 2.81(1.27–6.22) | 0.008 |
| Antidepressants | 41.4 | 17.4 | 3.36(2.39–4.74) | < 0.001 |
| Anxiolytics/Hypnotics | 10.1 | 7.6 | 1.36(0.79–2.36) | 0.168 |
| Respiratory and sensory drugs | 40.8 | 47.1 | 0.78(0.56–1.08) | 0.076 |

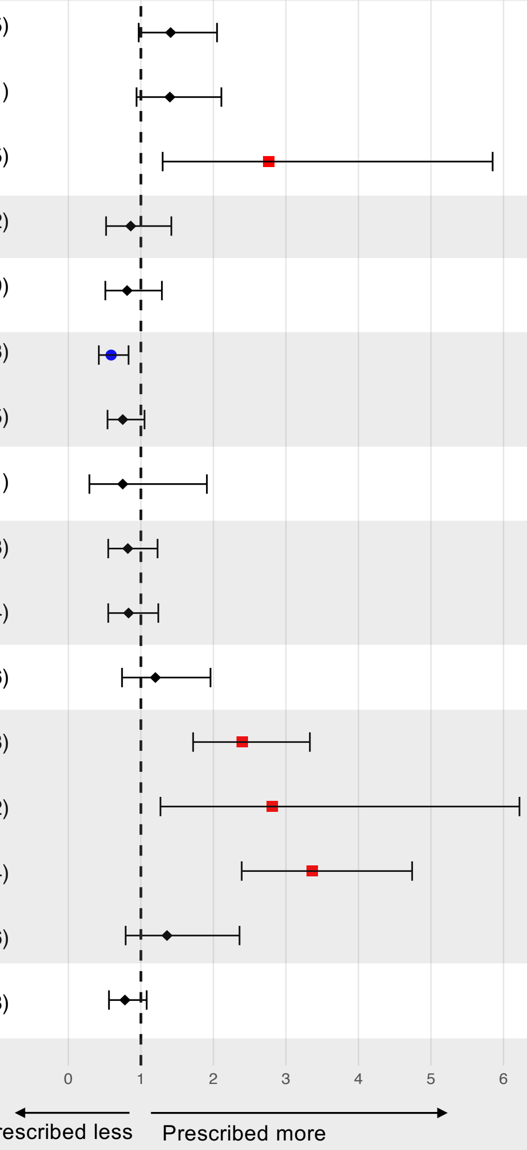

Supplementary Figure S4a.

### Comorbidity profiles of patients with severe anorexia nervosa (derived from latent class analysis)

Cluster 1: Severe AN patients without any comorbidities
N = 169, AN-RSI = 1.18 (1.17)[a]

Cluster 2: Severe AN patients with low burden of comorbidities
N = 459, AN-RSI = 1.27 (1.40)[a]

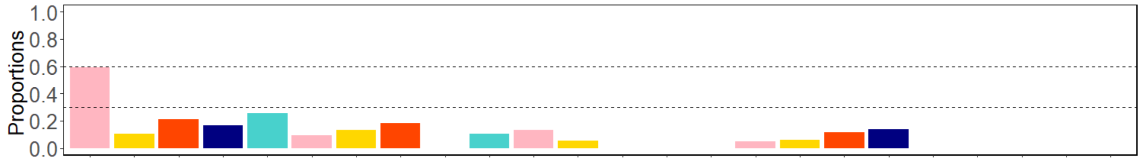


Cluster 3: Severe AN patients with medium burden of personality, mood, and anxiety disorders
N = 697, AN-RSI = 1.36 (1.17)[a]

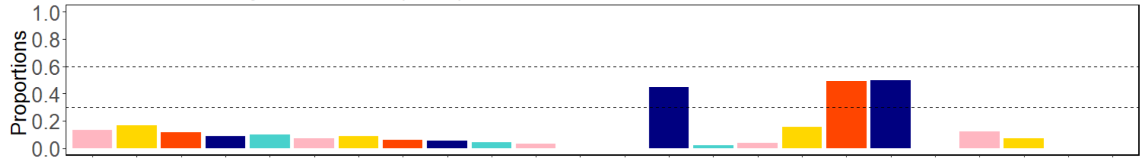


Cluster 4: Severe AN patients with high burden of personality disorders, and self-harm conditions
N = 168, AN-RSI = 1.83 (1.46)[a]

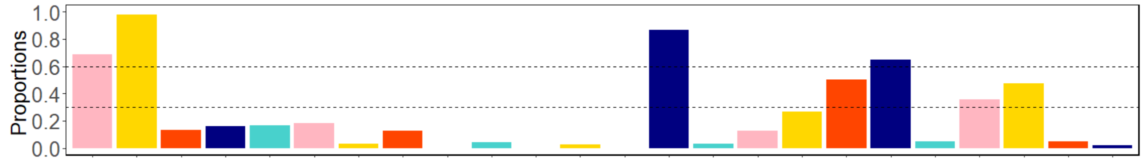


Cluster 5: Severe AN patients with high burden comorbidities
N = 159, AN-RSI = 1.94 (1.49)[a]

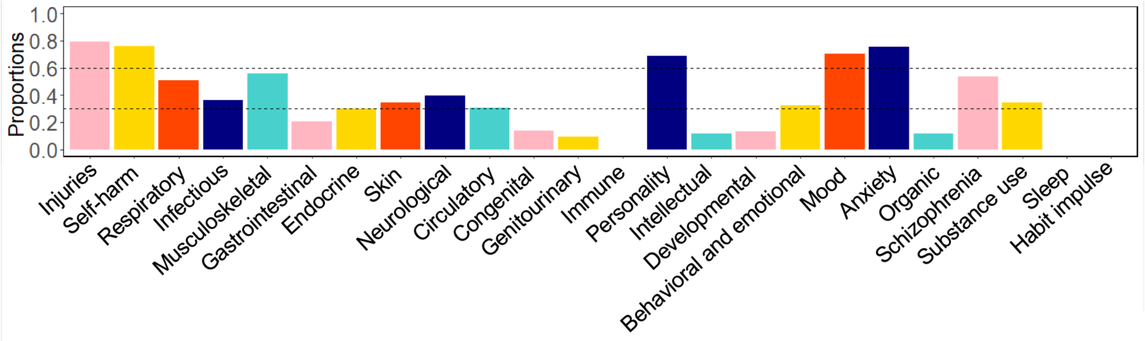


*Note.* AN-RSI: Anorexia Nervosa Register-based Severity Index
[a] Mean (standard deviation);
Comorbidities with less than five cases were set to zero.

Supplementary Figure S4b.

### Comorbidity profiles of patients with severe anorexia nervosa (derived from K-means)

Cluster 1: Severe AN patients without any comorbidities
N = 169, AN-RSI = 1.18 (1.17)[a]

Cluster 2: Severe AN patients with low burden of comorbidities
N = 619, AN-RSI = 1.22 (1.30)[a]

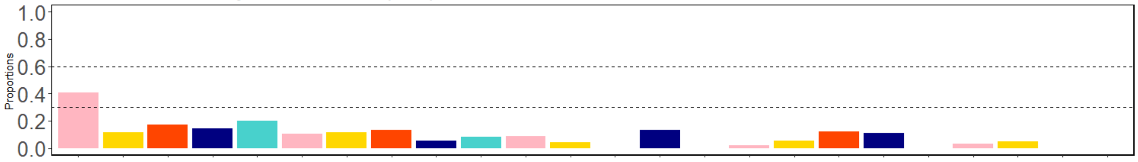


Cluster 3: Severe AN patients with medium burden of personality, mood, and anxiety disorders
N = 341, AN-RSI = 1.36 (1.27)[1]

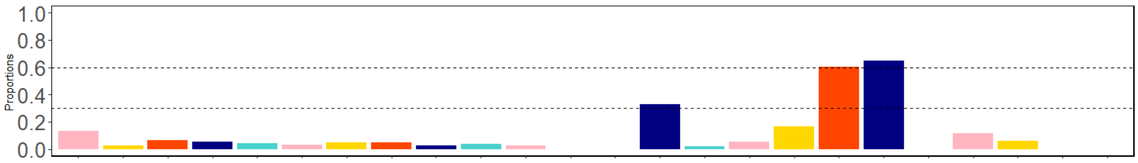


Cluster 4: Severe AN patients with high burden of personality disorders, and self-harm conditions
N = 369, AN-RSI = 1.64 (1.42)[a]

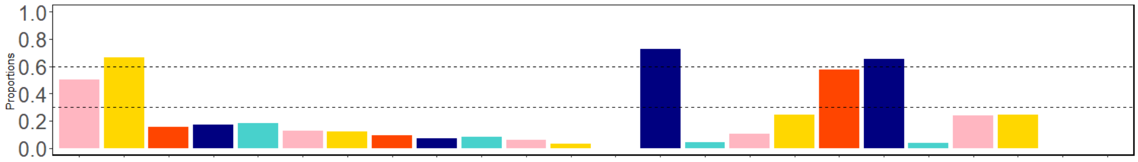


Cluster 5: Severe AN patients with high burden comorbidities
N = 154, AN-RSI = 1.81 (1.31)[a]

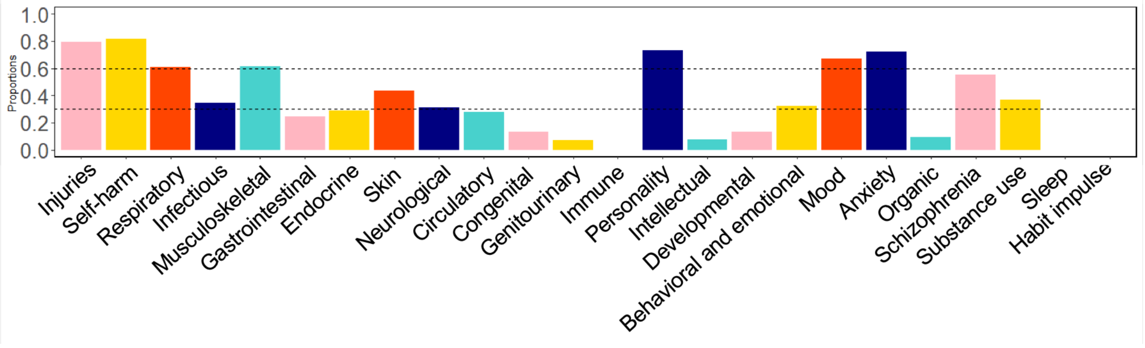


*Note.* AN-RSI: Anorexia Nervosa Register-based Severity Index
[a] Mean (standard deviation);
Comorbidities with less than five cases were set to zero.

Supplementary Figure S5.

### Medication prescriptions in severe anorexia nervosa patients with distinct comorbidity profiles

| Medication | Odds Ratio (95% CI) | P-value |
|---|---|---|
| **Cluster 5 vs 2** | | |
| Alimentary tract | 9.30(5.67–15.27) | <0.001 |
| Anti–infective drugs | 2.39(0.92–6.22) | 0.051 |
| Cardiovascular drugs | 3.06(2.10–4.46) | <0.001 |
| Genitourinary system and sex hormones | 1.13(0.75–1.69) | 0.322 |
| Hormonal preparation drugs | 2.57(1.63–4.07) | <0.001 |
| Immunomodulating drugs | 1.60(0.58–4.39) | 0.263 |
| Musculoskeletal drugs | 3.51(2.39–5.15) | <0.001 |
| Analgesics | 5.51(3.67–8.27) | <0.001 |
| Psychotropic drugs | 20.14(7.33–55.37) | <0.001 |
| Antidepressants | 7.82(4.73–12.93) | <0.001 |
| Antiepileptics | 23.97(13.89–41.37) | <0.001 |
| Anxiolytics/Hypnotics | 11.66(7.58–17.95) | <0.001 |
| Antipsychotics | 24.93(15.62–39.78) | <0.001 |
| Psychostimulants | 15.83(4.52–55.47) | <0.001 |
| Other NS drugs | 15.45(8.25–28.91) | <0.001 |
| Respiratory and sensory drugs | 3.36(2.12–5.32) | <0.001 |
| Other | 5.83(0.53–64.78) | 0.168 |
| **Cluster 4 vs 2** | | |
| Alimentary tract | 5.52(0.50–61.25) | 0.181 |
| Anti–infective drugs | 1.10(0.76–1.58) | 0.346 |
| Cardiovascular drugs | 7.42(3.91–14.09) | <0.001 |
| Genitourinary system and sex hormones | 1.53(1.07–2.19) | 0.013 |
| Hormonal preparation drugs | 0.49(0.11–2.24) | 0.267 |
| Immunomodulating drugs | 1.69(1.18–2.41) | 0.003 |
| Musculoskeletal drugs | 0.48(0.24–0.98) | 0.028 |
| Analgesics | 0.80(0.55–1.18) | 0.152 |
| Psychotropic drugs | 1.01(0.51–2.00) | 0.500 |
| Antidepressants | 1.28(0.87–1.90) | 0.127 |
| Antiepileptics | 2.69(1.86–3.88) | <0.001 |
| Anxiolytics/Hypnotics | 14.25(7.63–26.61) | <0.001 |
| Antipsychotics | 27.73(17.36–44.31) | <0.001 |
| Psychostimulants | 20.54(6.02–70.10) | <0.001 |
| Other NS drugs | 8.53(5.72–12.73) | <0.001 |
| Respiratory and sensory drugs | 11.57(6.68–20.05) | <0.001 |
| Other | 8.32(5.04–13.75) | <0.001 |
| **Cluster 3 vs 2** | | |
| Alimentary tract | 0.97(0.77–1.24) | 0.437 |
| Anti–infective drugs | 0.69(0.45–1.07) | 0.059 |
| Cardiovascular drugs | 0.92(0.70–1.21) | 0.291 |
| Genitourinary system and sex hormones | 0.86(0.67–1.12) | 0.150 |
| Hormonal preparation drugs | 0.67(0.45–0.99) | 0.029 |
| Immunomodulating drugs | 0.29(0.10–0.85) | 0.016 |
| Musculoskeletal drugs | 0.90(0.71–1.15) | 0.229 |
| Analgesics | 0.76(0.59–0.97) | 0.017 |
| Psychotropic drugs | 2.58(1.95–3.40) | <0.001 |
| Antidepressants | 3.02(2.36–3.87) | <0.001 |
| Antiepileptics | 3.51(2.13–5.76) | <0.001 |
| Anxiolytics/Hypnotics | 2.13(1.63–2.78) | <0.001 |
| Antipsychotics | 4.24(3.10–5.79) | <0.001 |
| Psychostimulants | 6.84(2.07–22.53) | <0.001 |
| Other NS drugs | 2.94(1.62–5.33) | <0.001 |
| Respiratory and sensory drugs | 0.75(0.59–0.95) | 0.010 |
| Other | 0.66(0.04–10.55) | 0.500 |

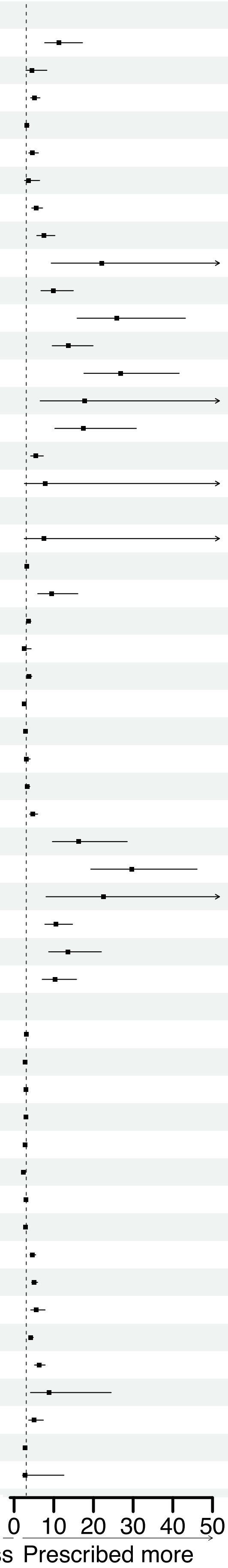


*Note.* CI: confidence interval
NSAID: nonsteroidal anti-inflammatory drugs
Other NS drugs: other nervous system drugs

Supplementary Figure S5.

### Medication prescriptions in severe anorexia nervosa patients with distinct comorbidity profiles (continued)

| Medication | Odds Ratio (95% CI) | P-value |
|---|---|---|
| **Cluster 5 vs 3** | | |
| Alimentary tract | 9.56(5.89–15.49) | <0.001 |
| Anti–infective drugs | 3.44(1.36–8.67) | 0.004 |
| Cardiovascular drugs | 3.34(2.34–4.78) | <0.001 |
| Genitourinary system and sex hormones | 1.30(0.88–1.92) | 0.109 |
| Hormonal preparation drugs | 3.85(2.45–6.04) | <0.001 |
| Immunomodulating drugs | 5.43(1.64–18.01) | 0.004 |
| Musculoskeletal drugs | 3.87(2.68–5.60) | <0.001 |
| Analgesics | 7.29(4.92–10.81) | <0.001 |
| Psychotropic drugs | 7.82(2.84–21.51) | <0.001 |
| Antidepressants | 2.59(1.57–4.25) | <0.001 |
| Antiepileptics | 6.84(4.68–9.98) | <0.001 |
| Anxiolytics/Hypnotics | 5.47(3.66–8.17) | <0.001 |
| Antipsychotics | 5.89(3.91–8.86) | <0.001 |
| Psychostimulants | 2.32(1.21–4.42) | 0.008 |
| Other NS drugs | 5.26(3.43–8.04) | <0.001 |
| Respiratory and sensory drugs | 4.49(2.88–7.02) | <0.001 |
| **Cluster 4 vs 3** | | |
| Alimentary tract | 2.76(1.94–3.92) | <0.001 |
| Anti–infective drugs | 1.45(0.77–2.74) | 0.157 |
| Cardiovascular drugs | 1.40(0.96–2.04) | 0.048 |
| Genitourinary system and sex hormones | 0.93(0.65–1.33) | 0.378 |
| Hormonal preparation drugs | 0.72(0.36–1.45) | 0.226 |
| Immunomodulating drugs | 1.67(0.32–8.67) | 0.446 |
| Musculoskeletal drugs | 1.86(1.33–2.62) | <0.001 |
| Analgesics | 2.02(1.43–2.86) | <0.001 |
| Psychotropic drugs | 2.88(1.51–5.48) | <0.001 |
| Antidepressants | 2.75(1.68–4.52) | <0.001 |
| Antiepileptics | 3.30(2.25–4.85) | <0.001 |
| Anxiolytics/Hypnotics | 4.00(2.77–5.78) | <0.001 |
| Antipsychotics | 6.55(4.35–9.87) | <0.001 |
| Psychostimulants | 3.00(1.66–5.44) | <0.001 |
| Other NS drugs | 4.85(3.18–7.39) | <0.001 |
| Respiratory and sensory drugs | 1.46(1.04–2.07) | 0.019 |
| Other | 8.39(0.76–93.03) | 0.090 |
| **Cluster 5 vs 4** | | |
| Alimentary tract | 3.46(1.98–6.06) | <0.001 |
| Anti–infective drugs | 2.37(0.82–6.88) | 0.084 |
| Cardiovascular drugs | 2.39(1.52–3.76) | <0.001 |
| Genitourinary system and sex hormones | 1.40(0.87–2.26) | 0.104 |
| Hormonal preparation drugs | 5.31(2.55–11.05) | <0.001 |
| Immunomodulating drugs | 3.25(0.65–16.37) | 0.124 |
| Musculoskeletal drugs | 2.08(1.32–3.26) | |
| Analgesics | 3.60(2.25–5.76) | <0.001 |
| Psychotropic drugs | 2.71(0.85–8.71) | 0.070 |
| Antidepressants | 0.94(0.48–1.82) | 0.493 |
| Antiepileptics | 2.07(1.33–3.23) | <0.001 |
| Anxiolytics/Hypnotics | 1.37(0.83–2.25) | 0.136 |
| Antipsychotics | 0.90(0.53–1.54) | 0.400 |
| Psychostimulants | 0.77(0.38–1.56) | 0.293 |
| Other NS drugs | 1.08(0.68–1.73) | 0.412 |
| Respiratory and sensory drugs | 3.07(1.82–5.18) | <0.001 |
| Other | 1.06(0.15–7.60) | 0.500 |

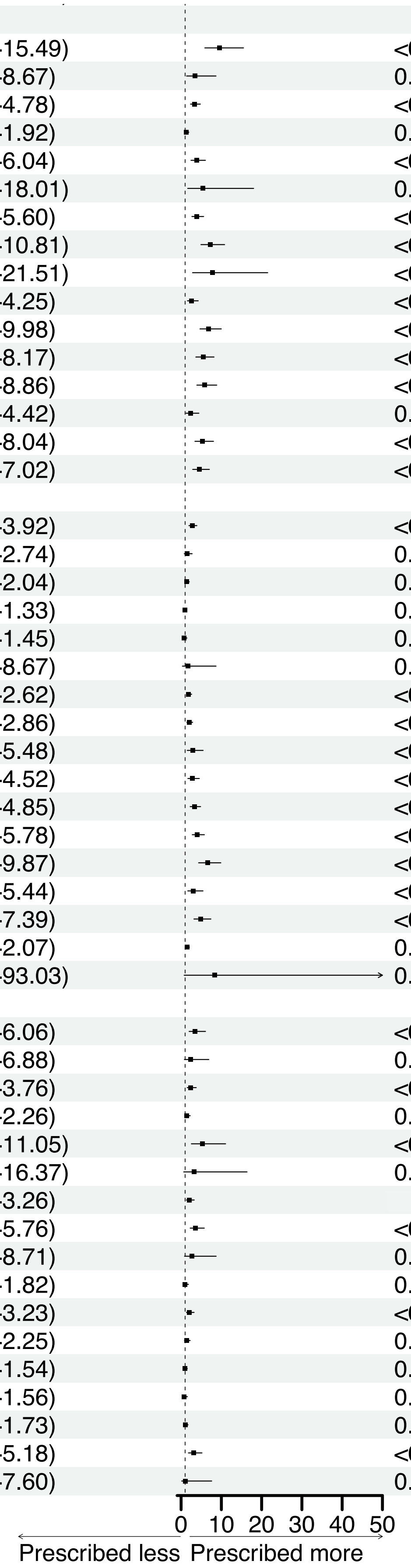


*Note.* CI: confidence interval
NSAID: nonsteroidal anti-inflammatory drugs
Other NS drugs: other nervous system drugs

Supplementary Figure S6.

### Medication prescriptions in patients with severe vs less-severe anorexia nervosa between 5 and 10 years post-diagnosis (severity evaluated at 3 years post-diagnosis)

| Medication | Severe AN (%) | Less–severe AN (%) | Odds Ratio (95% CI) | P–value |
|---|---|---|---|---|
| Alimentary tract | 36.3 | 30.5 | 1.30(1.16–1.46) | < 0.001 |
| Gastrointestinal | 28.9 | 25.3 | 1.20(1.06–1.36) | 0.002 |
| Vitamins | 0.7 | 0.5 | 1.36(0.68–2.72) | 0.251 |
| Minerals | 5.0 | 3.1 | 3.05(2.44–3.81) | < 0.001 |
| Cardiovascular drugs | 21.2 | 20.6 | 1.04(0.91–1.19) | 0.301 |
| Iron preparations | 1.6 | 1.2 | 1.33(0.85–2.10) | 0.131 |
| Anti–infective drugs | 65.0 | 68.1 | 0.80(0.69–0.94) | 0.004 |
| Genitourinary system and sex hormones | 65.4 | 70.5 | 0.79(0.70–0.89) | < 0.001 |
| Sex hormones | 64.2 | 68.6 | 0.82(0.73–0.92) | < 0.001 |
| Hormonal preparation drugs | 8.3 | 7.7 | 1.09(0.90–1.33) | 0.203 |
| Thyroid therapy | 3.2 | 2.8 | 1.15(0.84–1.58) | 0.211 |
| Musculoskeletal drugs | 30.7 | 34.7 | 0.83(0.74–0.94) | 0.001 |
| NSAID | 29.5 | 33.8 | 0.82(0.73–0.92) | < 0.001 |
| Biphosphonates | 1.1 | 0.5 | 2.02(1.13–3.61) | 0.012 |
| Immunomodulating drugs | 1.1 | 1.6 | 0.67(0.40–1.11) | 0.071 |
| Analgesics | 30.3 | 27.6 | 1.14(1.02–1.29) | 0.015 |
| Psychotropic drugs | 64.7 | 52.8 | 1.63(1.46–1.83) | < 0.001 |
| Antiepileptics | 13.1 | 6.8 | 1.55(1.31–1.83) | < 0.001 |
| Antipsychotics | 24.4 | 13.8 | 2.01(1.76–2.30) | < 0.001 |
| Antidepressants | 47.4 | 35.0 | 1.67(1.50–1.87) | < 0.001 |
| Anxiolytics/Hypnotics | 28.5 | 20.3 | 1.56(1.38–1.77) | < 0.001 |
| Psychostimulants | 3.4 | 3.5 | 0.96(0.71–1.29) | 0.413 |
| Other NS drugs | 6.6 | 5.4 | 1.66(1.35–2.04) | < 0.001 |
| Respiratory and sensory drugs | 45.2 | 47.9 | 0.90(0.80–1.00) | 0.025 |
| Other | 0.4 | 0.4 | 0.96(0.42–2.22) | 0.500 |

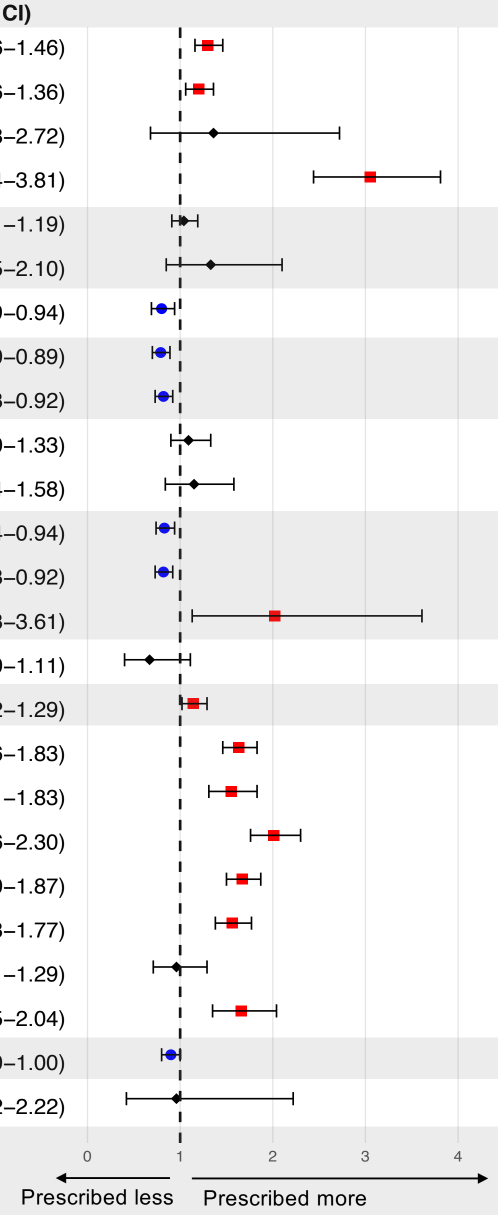

Supplementary Figure S7.

### Medication prescriptions in patients with severe vs less-severe anorexia nervosa between 7 and 10 years post-diagnosis (severity evaluated at 7 years post-diagnosis)

| Medication | Severe AN (%) | Less–severe AN (%) | Odds Ratio (95% CI) | P–value |
|---|---|---|---|---|
| Alimentary tract | 53.9 | 23.2 | 1.70(1.51–1.91) | < 0.001 |
| Gastrointestinal | 25.2 | 18.6 | 1.48(1.30–1.68) | < 0.001 |
| Vitamins | 0.5 | 0.3 | 1.56(0.68–3.59) | 0.208 |
| Minerals | 8.8 | 2.0 | 4.76(3.73–6.09) | < 0.001 |
| Cardiovascular drugs | 19.4 | 15.9 | 1.27(1.10–1.45) | < 0.001 |
| Iron preparations | 1.4 | 0.7 | 2.01(1.22–3.33) | 0.004 |
| Anti–infective drugs | 76.6 | 79.3 | 0.85(0.75–0.97) | 0.009 |
| Genitourinary system and sex hormones | 55.6 | 62.7 | 0.75(0.67–0.83) | < 0.001 |
| Sex hormones | 54.1 | 60.8 | 0.76(0.68–0.85) | < 0.001 |
| Hormonal preparation drugs | 7.5 | 6.2 | 1.23(1.00–1.51) | 0.030 |
| Thyroid therapy | 3.1 | 2.5 | 1.24(0.90–1.71) | 0.104 |
| Musculoskeletal drugs | 26.0 | 26.5 | 0.97(0.86–1.10) | 0.350 |
| NSAID | 24.5 | 25.5 | 0.95(0.84–1.08) | 0.232 |
| Biphosphonates | 1.2 | 0.3 | 4.15(2.17–7.94) | < 0.001 |
| Immunomodulating drugs | 0.9 | 1.2 | 0.75(0.43–1.31) | 0.187 |
| Analgesics | 25.5 | 22.1 | 1.20(1.06–1.36) | 0.002 |
| Psychotropic drugs | 61.7 | 45.0 | 1.97(1.76–2.20) | < 0.001 |
| Antiepileptics | 12.6 | 7.0 | 1.92(1.62–2.29) | < 0.001 |
| Antipsychotics | 23.2 | 10.0 | 2.72(2.37–3.13) | < 0.001 |
| Antidepressants | 45.1 | 36.7 | 2.26(2.03–2.53) | < 0.001 |
| Anxiolytics/Hypnotics | 25.0 | 14.6 | 1.95(1.71–2.22) | < 0.001 |
| Psychostimulants | 3.6 | 2.7 | 1.36(1.01–1.83) | 0.026 |
| Other NS drugs | 7.6 | 4.1 | 1.93(1.55–2.40) | < 0.001 |
| Respiratory and sensory drugs | 36.9 | 38.7 | 0.93(0.83–1.04) | 0.096 |
| Other | 0.2 | 0.4 | 0.64(0.22–1.85) | 0.273 |

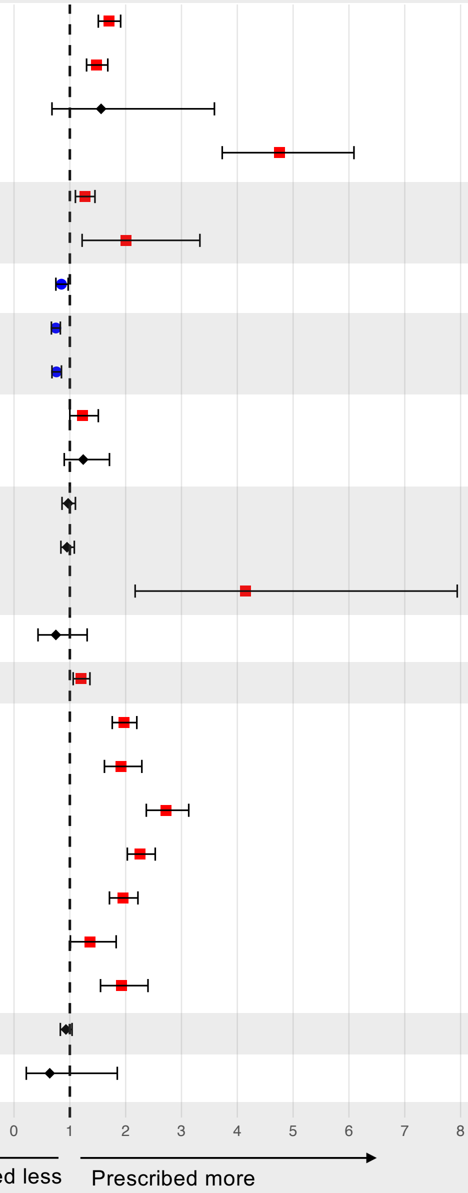

Supplementary Figure S8.

### Comorbidity profiles of patients with severe anorexia nervosa (severity evaluated at 3 years post-diagnosis, derived from latent class analysis)

**Cluster 1: Severe AN patients without any comorbidities**

N = 179, AN-RSI = 1.13 (1.15)[a]

**Cluster 2: Severe AN patients with low burden of comorbidities**

N = 571, AN-RSI = 1.16 (1.25)[a]

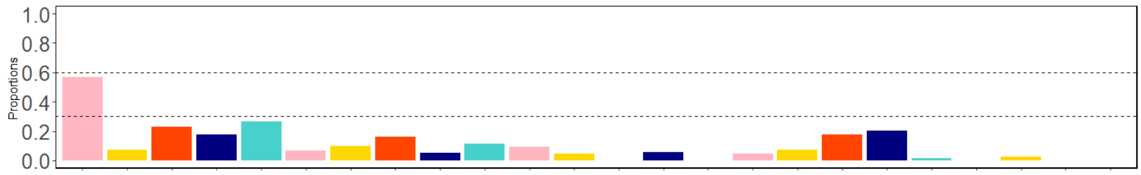


**Cluster 3: Severe AN patients with medium burden of personality, mood, and anxiety disorders**

N = 493, AN-RSI = 1.29 (1.07)[a]

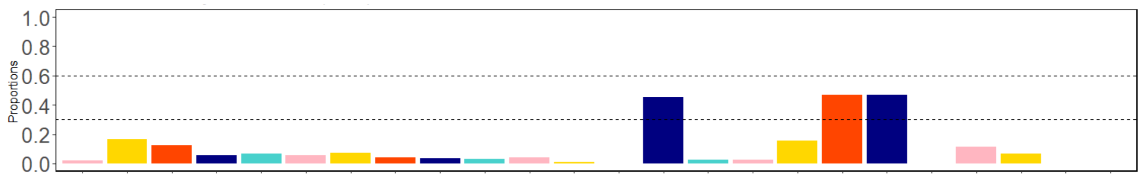


**Cluster 4: Severe AN patients with high burden of personality disorders, and self-harm conditions**

N = 298, AN-RSI = 1.64 (0.95)[a]

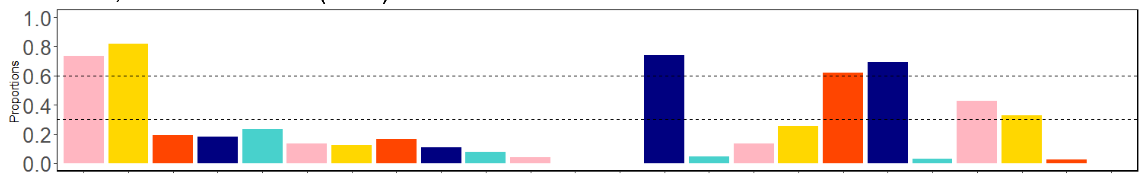


**Cluster 5: Severe AN patients with high burden comorbidities**

N = 77, AN-RSI = 1.54 (1.27)[a]

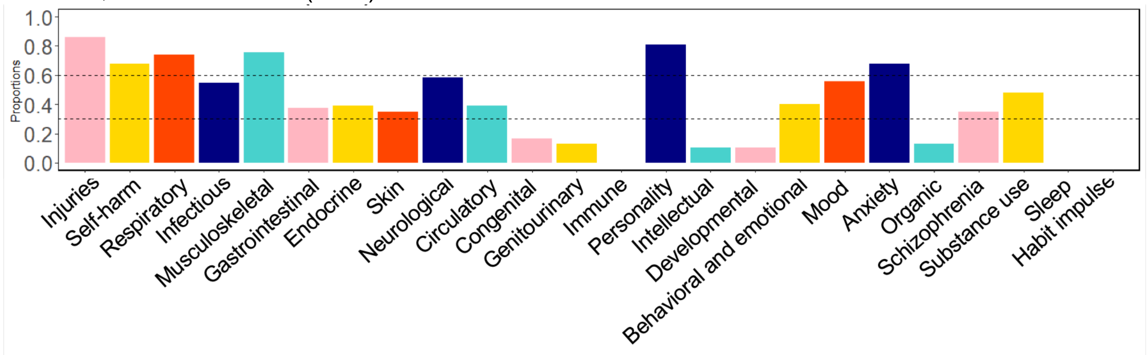


*Note.* AN-RSI: Anorexia Nervosa Register-based Severity Index
[a] Mean (standard deviation);
Comorbidities with less than five cases were set to zero.

Supplementary Figure S9.

### Comorbidity profiles of patients with severe anorexia nervosa (severity evaluated at 7 years post-diagnosis, derived from latent class analysis)

**Cluster 1: Severe AN patients without any comorbidities**

N = 179, AN-RSI = 1.13 (1.15)[a]

**Cluster 2: Severe AN patients with low burden of comorbidities**

N = 456, AN-RSI = 1.21 (1.45)[a]

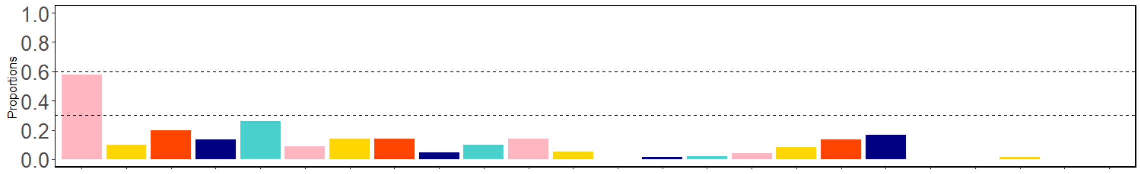


**Cluster 3: Severe AN patients with medium burden of personality, mood, and anxiety disorders**

N = 726, AN-RSI = 1.31 (1.39)[a]

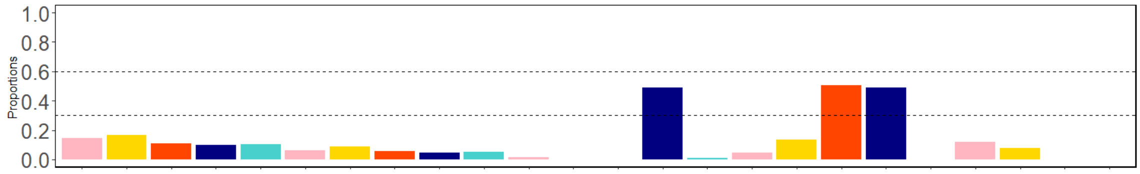


**Cluster 4: Severe AN patients with high burden of personality disorders, and self-harm conditions**

N = 257, AN-RSI = 1.50 (1.34)[a]

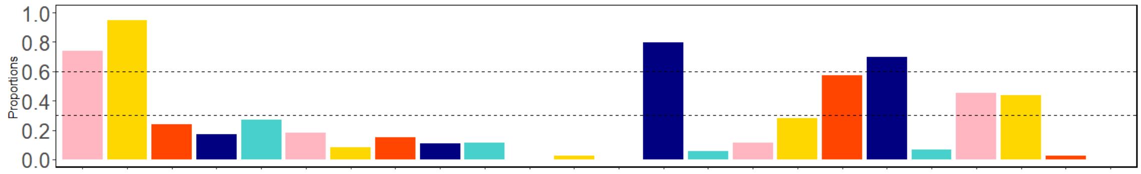


**Cluster 5: Severe AN patients with high burden comorbidities**

N = 101, AN-RSI = 1.86 (1.64)[a]

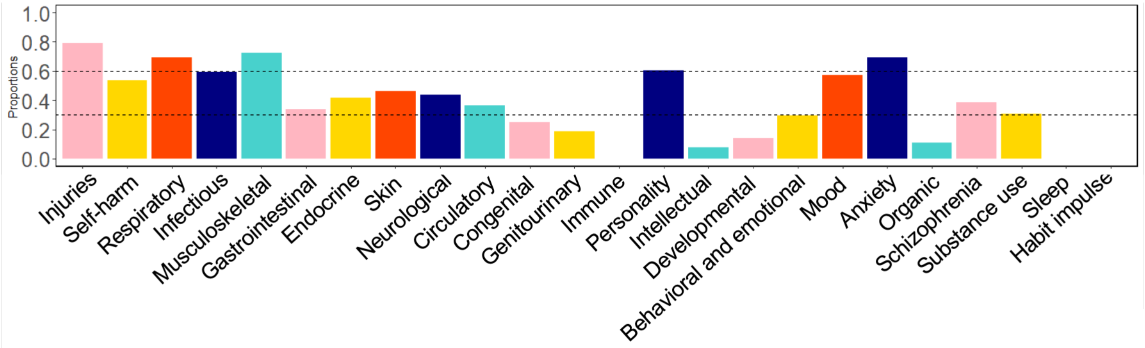


*Note.* AN-RSI: Anorexia Nervosa Register-based Severity Index
[a] Mean (standard deviation);
Comorbidities with less than five cases were set to zero.
